# A scoping review of lesbian, gay, bisexual, transgender, queer, and intersex (LGBTQI+) people’s health in India

**DOI:** 10.1101/2022.11.16.22282390

**Authors:** Venkatesan Chakrapani, Peter A. Newman, Murali Shunmugam, Shruta Rawat, Biji R. Mohan, Dicky Baruah, Suchon Tepjan

**Affiliations:** Centre for Sexuality and Health Research and Policy (C-SHaRP), Chennai, India; The Humsafar Trust, Mumbai, India; Factor-Inwentash Faculty of Social Work, University of Toronto, Toronto, Ontario, Canada; VOICES-Thailand Foundation, Chiang Mai, Thailand

**Keywords:** LGBTQI persons, Sexual and gender minorities, Social determinants of health, Health inequalities, HIV, Mental health, Scoping review, India

## Abstract

Amid incremental progress in establishing an enabling legal and policy environment for lesbian, gay, bisexual, transgender and queer-identified people, and people with intersex variations (LGBTQI+) in India, evidence gaps on LGBTQI+ health are of increasing concern. To that end, we conducted a scoping review to map and synthesize the current evidence base, identify research gaps, and provide recommendations for future research. We conducted a scoping review using the Joanna Briggs Institute methodology. We systematically searched 14 databases to identify peer-reviewed journal articles published in English language between January 1, 2010 and November 20, 2021, that reported empirical qualitative, quantitative or mixed methods data on LGBTQI+ people’s health in India. Out of 3,003 results in total, we identified 177 eligible articles; 62% used quantitative, 31% qualitative, and 7% mixed methods. The majority (55%) focused on gay and other men who have sex with men (MSM), 16% transgender women, and 14% both of these populations; 4% focused on lesbian and bisexual women, and 2% on transmasculine people. Overall, studies reported high HIV and sexually transmitted infection prevalence; multilevel risk factors for HIV; high levels of mental health burden linked to stigma, discrimination and violence victimization; and non-availability of gender-affirmative medical services in government hospitals. Few longitudinal studies and intervention studies were identified. Overall, LGBTQI+ health research in India needs to move beyond the predominant focus on HIV, and gay men/MSM and transgender women, to include mental health and non-communicable diseases, and individuals across the LGBTQI+ spectrum. Future research should build on largely descriptive studies to include explanatory and intervention studies, beyond urban to rural sites, and examine healthcare and service needs among LGBTQI+ people across the life course. Dedicated funding and training for junior investigators conducting LGBTQI+ health research is crucial to building a comprehensive evidence base to inform health policies and programs.

## INTRODUCTION

The right to the highest attainable standard of health is both universal and fundamental in international law [1]. This is enshrined in Article 12 [2] of the *Convention on Social, Economic, and Cultural Rights* and underlies United Nations Sustainable Development Goal 3 (SDG-3), which promises “Health for All” by 2030 and that “no one will be left behind.” This includes lesbian, gay, bisexual, transgender, queer identified, and people with intersex variations (LGBTQI+), who are entitled to the same standard of health as everyone else [3].

Despite the promise of the SDGs, evidence from across the globe suggests that LGBTQI+ health consistently lags behind that of the general public. Systematic and scoping reviews on health and healthcare access among LGBTQI+ people in high-income countries have shown that they continue to face disproportionate physical and mental health burdens [4–9]. For example, global reviews and large-scale studies have documented high levels of problematic alcohol use [10], sexualized drug use [11], mental health problems [4, 12], and high rates of HIV and other sexually transmitted infections (STIs) [13–15] among various LGBTQI+ subpopulations. Consistent with the minority stress model [16], many of these poor health outcomes are associated with societal stigma, discrimination, and violence, and systemic barriers in access to health services experienced by LGBTQI+ individuals [9, 17, 18].

Increasing recognition of health issues and disparities faced by LGBTQI+ people in the context of advances in LGBTQI+ rights movements globally have contributed to an evolving legal and policy environment that is gradually becoming more supportive of LGBTQI+ rights, and more attuned to addressing LGBTQI+ health disparities and discrimination [19]. These advances in the recognition of LGBTQI+ rights have concomitantly contributed to increasing awareness of the need for research evidence to meaningfully implement this policy shift. Population-specific data are sorely needed to document gaps, disparities, and progress in LGBTQI+ health over time, as recognized by numerous bodies including the World Bank and UNDP; both have called for more attention and investment in research on LGBTQI+ health [20]. This trend is evident in India where the decriminalization of adult consensual same-sex relationships (2018) [21] and the enactment of the Transgender Persons Protection of Rights Act (2019) [22] have recently emerged in rapid succession. The latter act was designed, among other things, to support and promote the delivery of non-discriminatory and gender-affirmative health services to transgender people. Subsequently, India’s Ministry of Social Justice and Empowerment’s expert committee on issues related to transgender persons has called for research evidence to design interventions to improve the health of transgender people [23].

We are aware of no overview and thorough mapping of the evidence base on LGBTQI+ health in India. A few published reviews of LGBTQI+ health in India have focused on specific topics, such as HIV-research among MSM or mental health issues among LGBTQI+ individuals [24–26]. To address the fragmented nature of current research knowledge, we conducted a scoping review to synthesize the evidence on LGBTQI+ health in India. The aim of this review was to characterize the breadth of published research on LGBTQI+ health in India and identify gaps in the evidence base, to provide recommendations for future research, and to better position existing evidence to inform health policies and interventions to advance LGBTQI+ health.

## METHODS

We used the scoping review framework initially proposed by Arksey and O’Malley [27] and advanced by the Joanna Briggs Institute [28]. The key steps involved: (1) identifying the research questions; (2) identifying relevant studies; (3) study selection using a pre-defined set of inclusion and exclusion criteria; (4) charting the data; and (5) collating, summarizing and reporting the results.

### Research questions

The specific questions guiding this review were: (1) What are the peer-reviewed literature sources available on LGBTQI+ health in India?; (2) What health problems and conditions are reported among LGBTQI+ people?; and (3) What are the gaps in the available evidence on LGBTQI+ health in India?

### Identifying studies from academic databases

A literature search was conducted using the following academic databases: Medline, Education Resources Information Centre (ERIC), Applied Social Sciences Index and Abstracts (ASSIA), Public Affairs Information Service Index (PAIS Index), Bibliography of Asian Studies, EconLit, Education Source, Social Work Abstracts, Sociological Abstracts, PsychInfo, LGBTLife, Gender Studies, HeinOnline, ProQuest Thesis, Worldwide Political Science Abstracts, and Child and Adolescent Development. Search strings previously validated for LGBT+ populations [29] were used for identifying relevant articles. Search strings were customized to account for the unique syntax of each database surveyed (see S1 Appendix). We added relevant Indian LGBT+ terminology, including indigenous sexual role-based identity terms such as kothi (feminine same-sex attracted males, primarily receptive sexual role), panthi (masculine and insertive role) and double-decker (both insertive and receptive role), and indigenous trans identities such as hijras. Given the absence of commonly-used Indian language terms for transgender men or transmasculine individuals, English language terms such as transgender men and transmasculine persons were used. We also added the term “India*” to the search string to limit the results geographically. The searches from each database were documented, duplicates were eliminated, and citations were imported to Covidence (Veritas Health Innovation, Melbourne) for abstract and full-text screening.

### Study Selection

Studies were selected according to pre-defined inclusion criteria. Studies must have been: 1) published between January 1, 2010 and November 20, 2021; conducted among LGBTQI+ people in India; 3) written in English; 4) peer reviewed; and 5) report primary data (qualitative, quantitative, or mixed methods). Two independent reviewers first screened the titles and abstracts for inclusion. In the case of discrepancies, a third reviewer was consulted to reach consensus. Full texts of potentially relevant articles were screened using a similar process.

### Charting, collating and summarizing the results

The following data were extracted for analysis: year of publication, study location, sample size, study population, objectives, design, methodology (qualitative, quantitative or mixed methods) and key findings. We summarized the results using frequencies, and thematic analysis and synthesis [28]. Studies were grouped by key themes that emerged from the synthesis: prevalence of HIV and STIs, and risk factors; stigma, discrimination and violence, and health impact; access to health services; interventions to improve health outcomes among LGBTQI+ populations; new HIV prevention technologies and their acceptability; and under-represented LGBTQI+ populations.

## RESULTS

### Study selection

The search strategy yielded 2,326 sources after removing duplicates. Screening of the titles and abstracts yielded 588 articles included in full-text review. Of these, 177 peer-reviewed articles met the *a priori* eligibility criteria and were included in the scoping review. The study selection process is shown in Figure 1. A summary of study characteristics and key findings are presented in Table 1.

**Fig 1.**
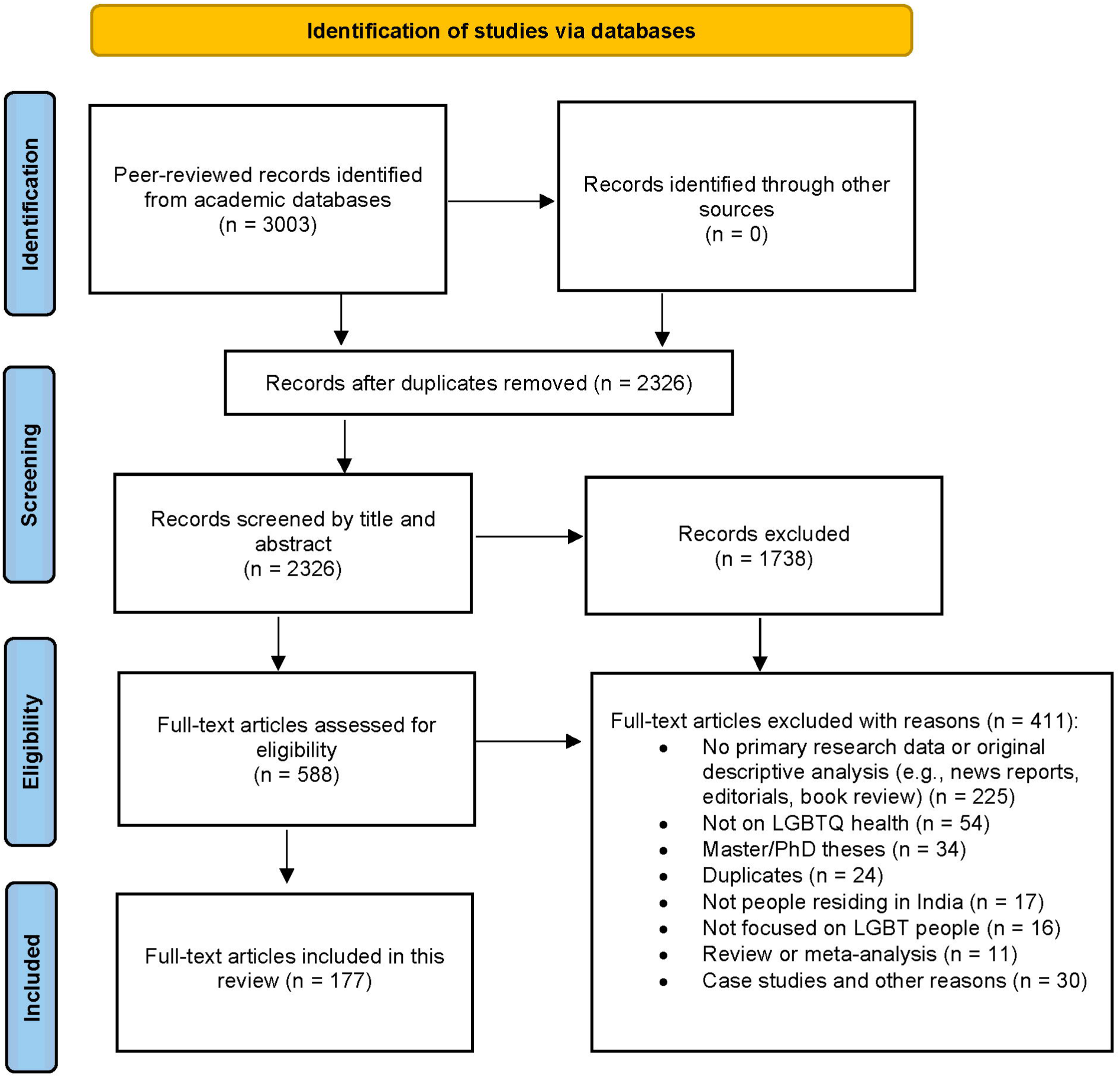
PRISMA Flowchart of study selection

**Table 1.**
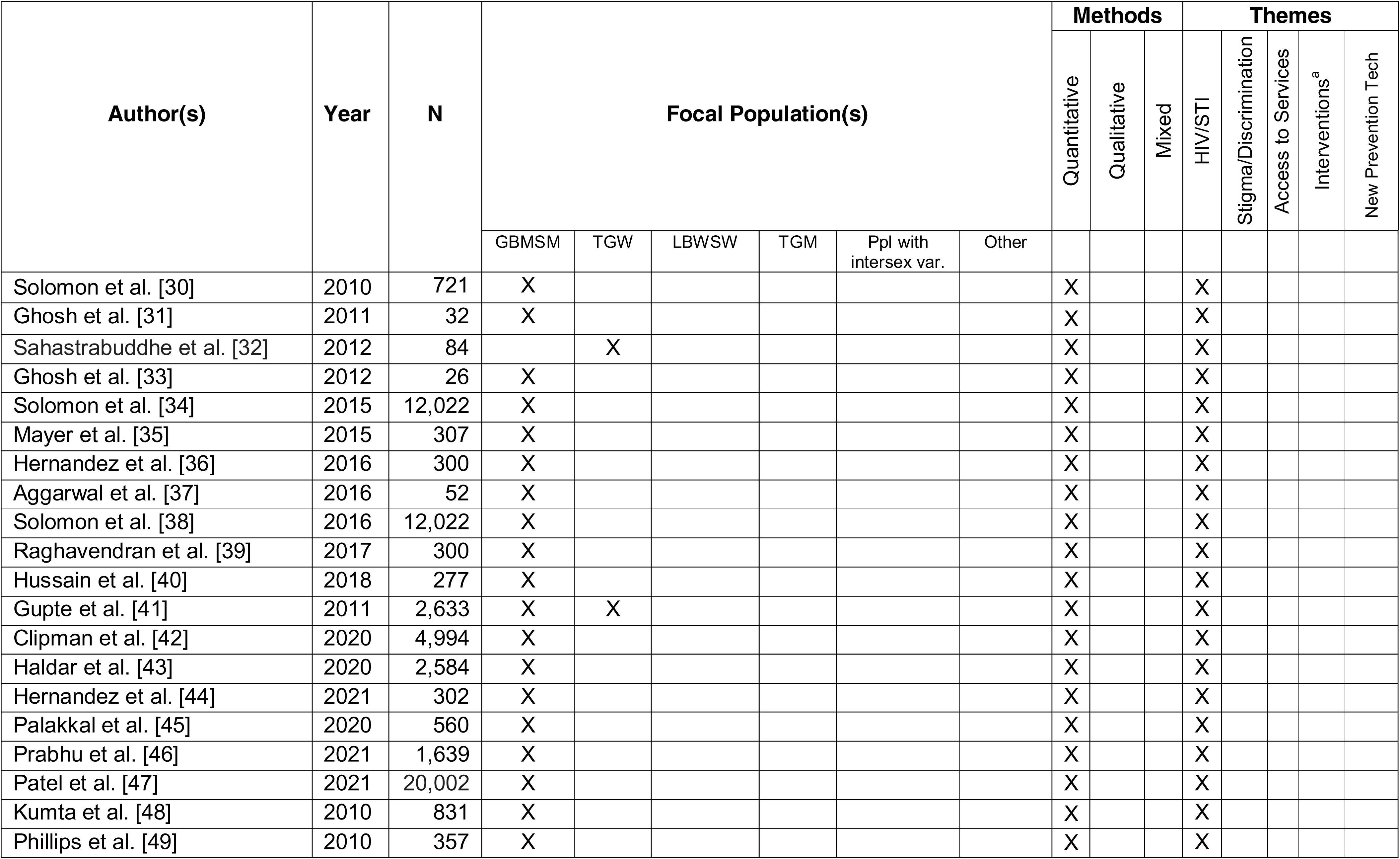
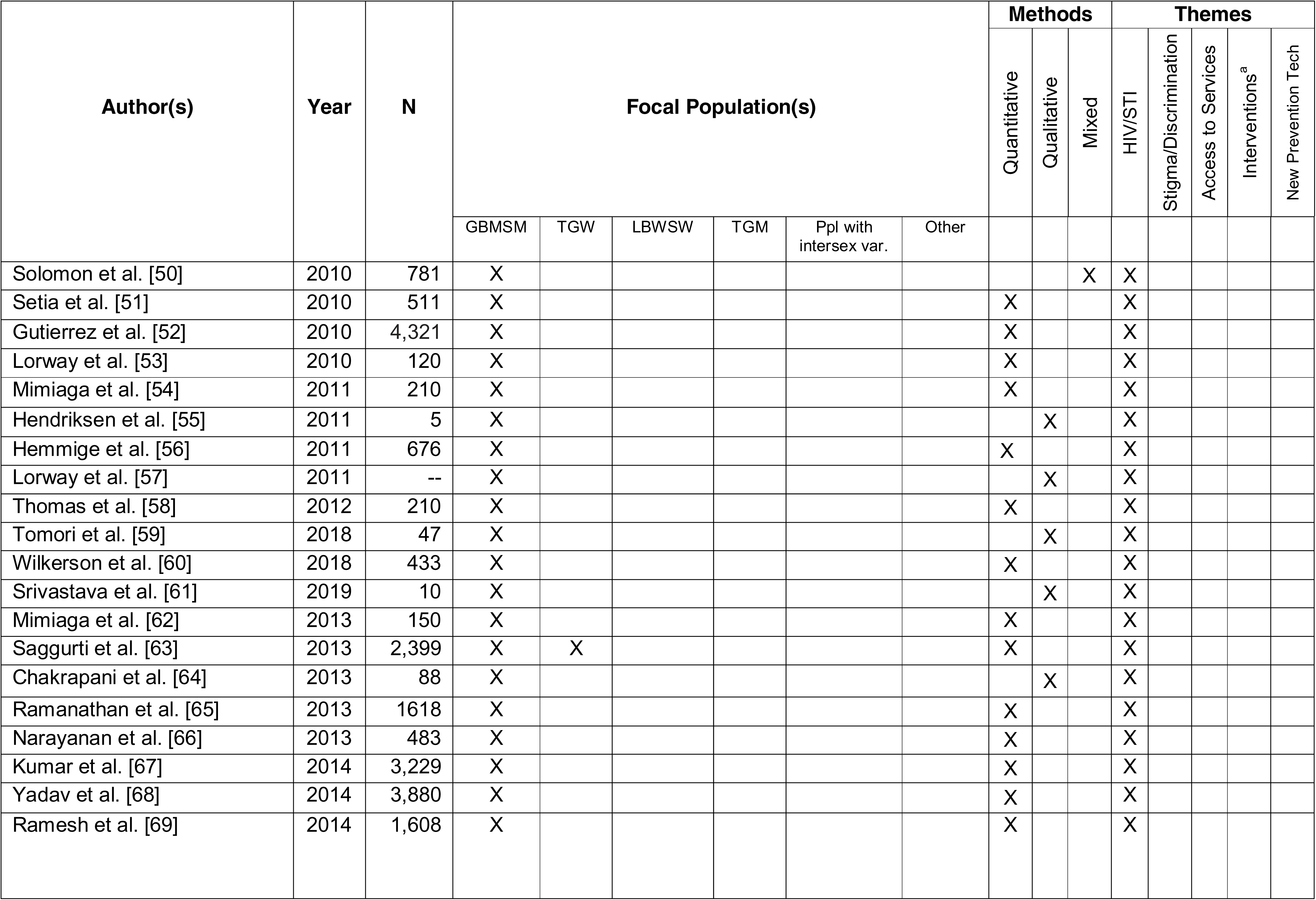
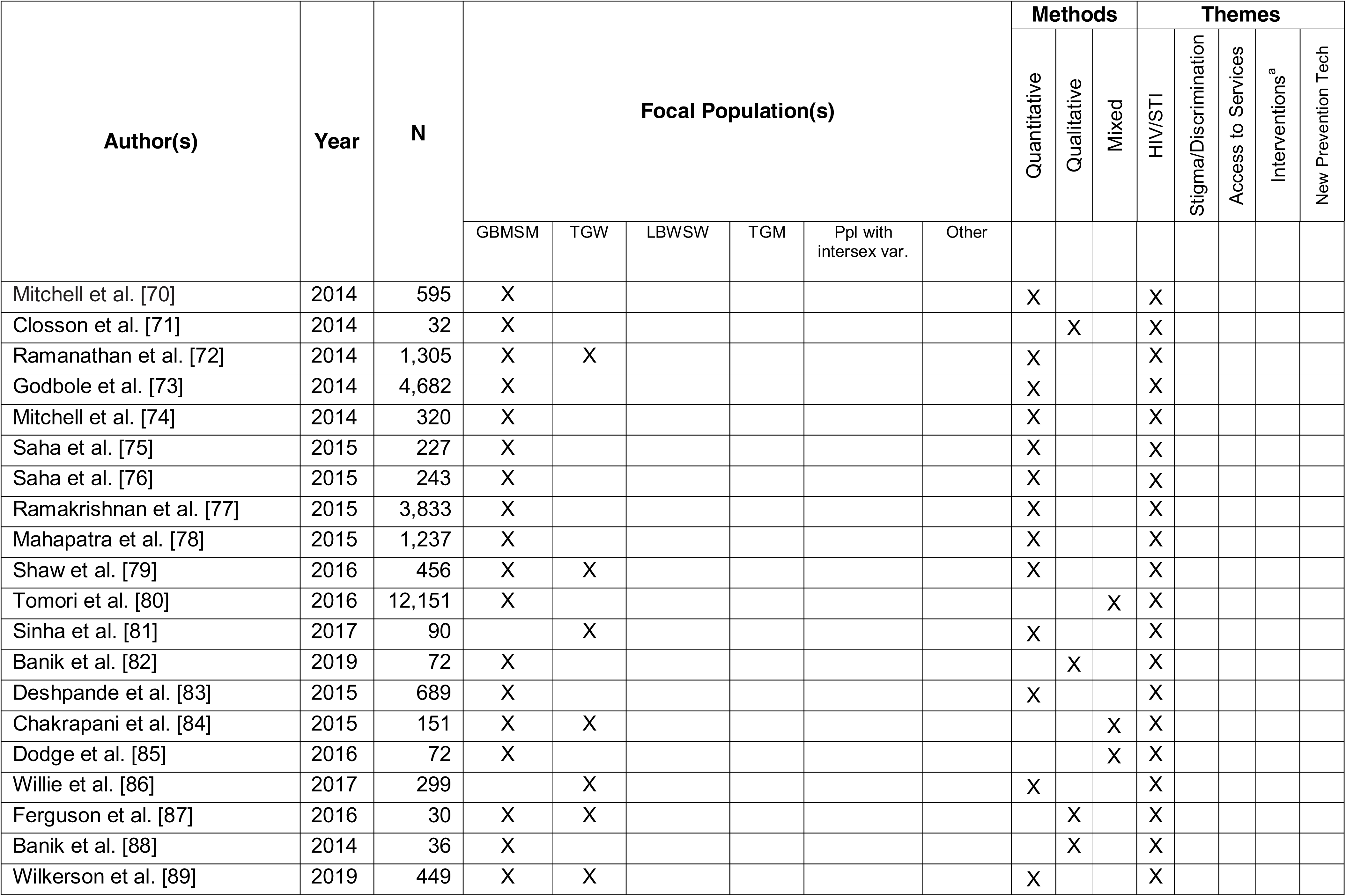
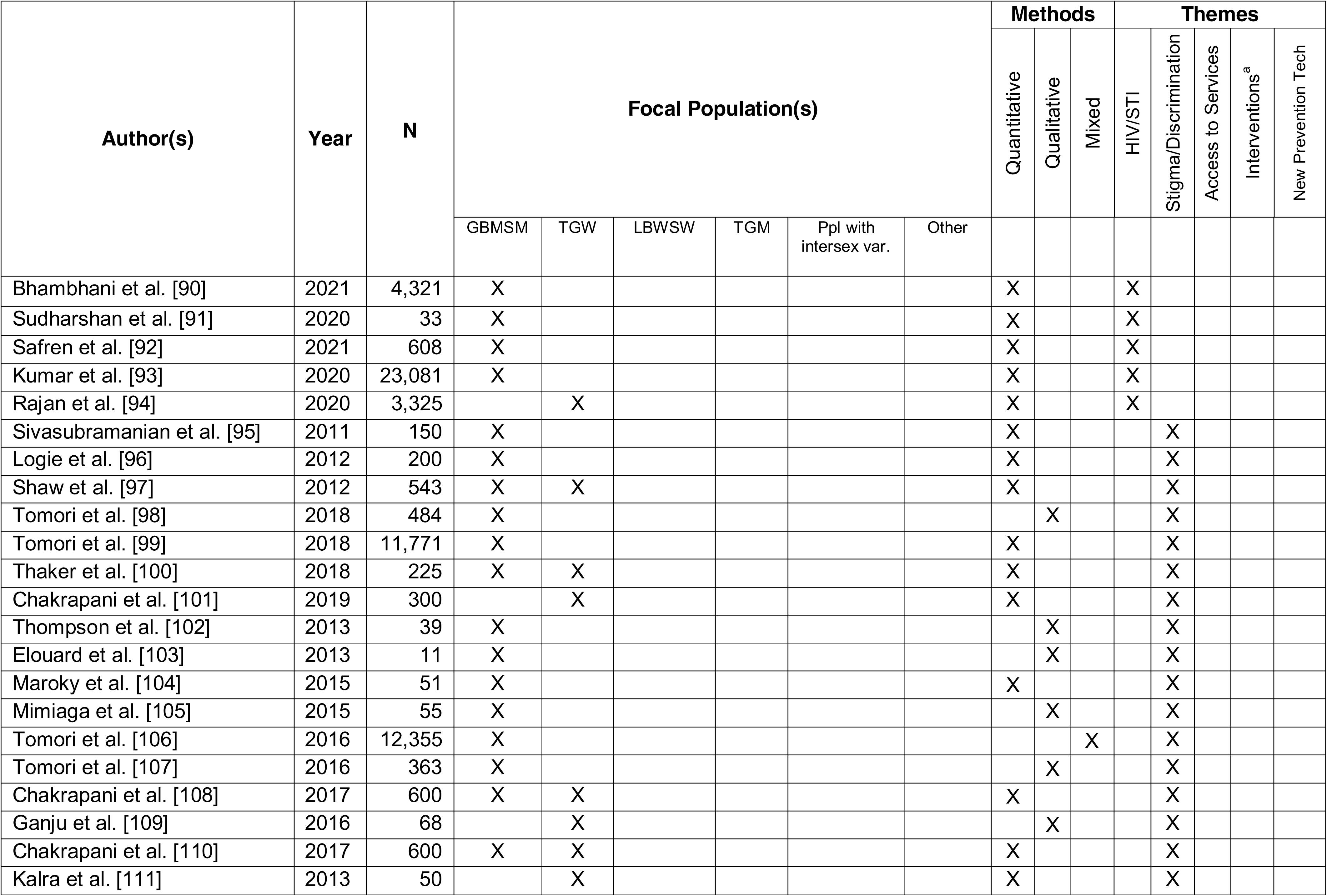
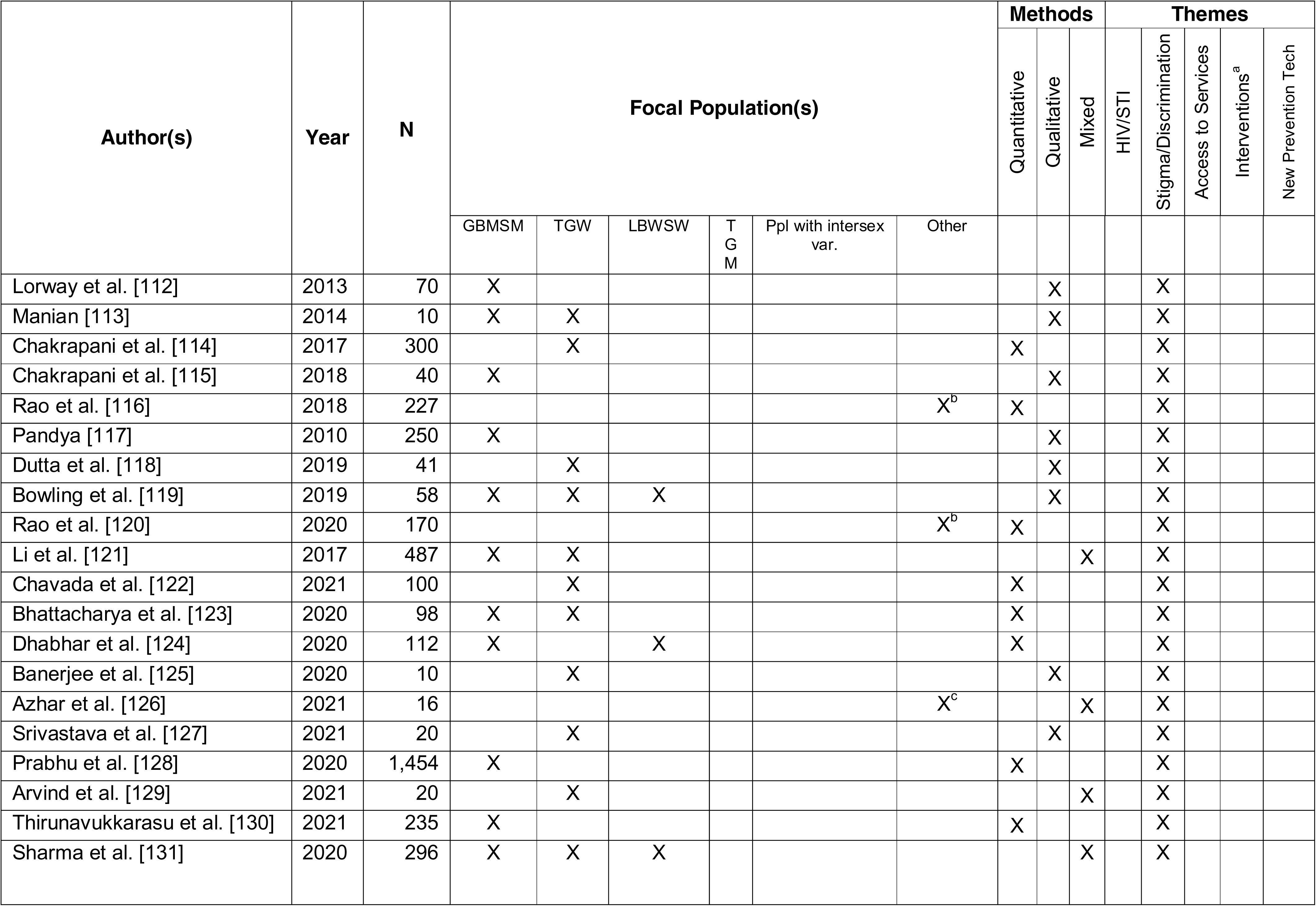
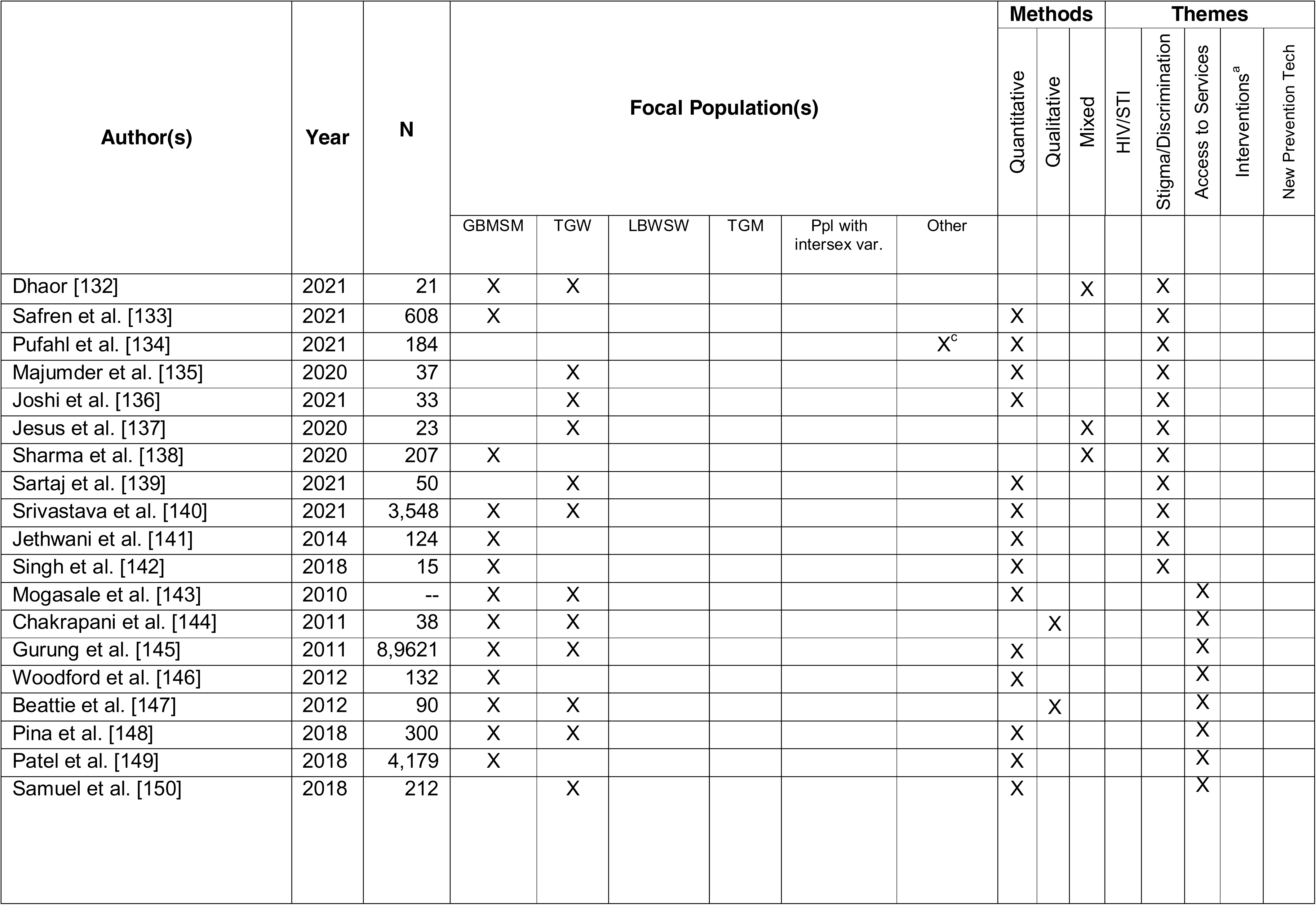
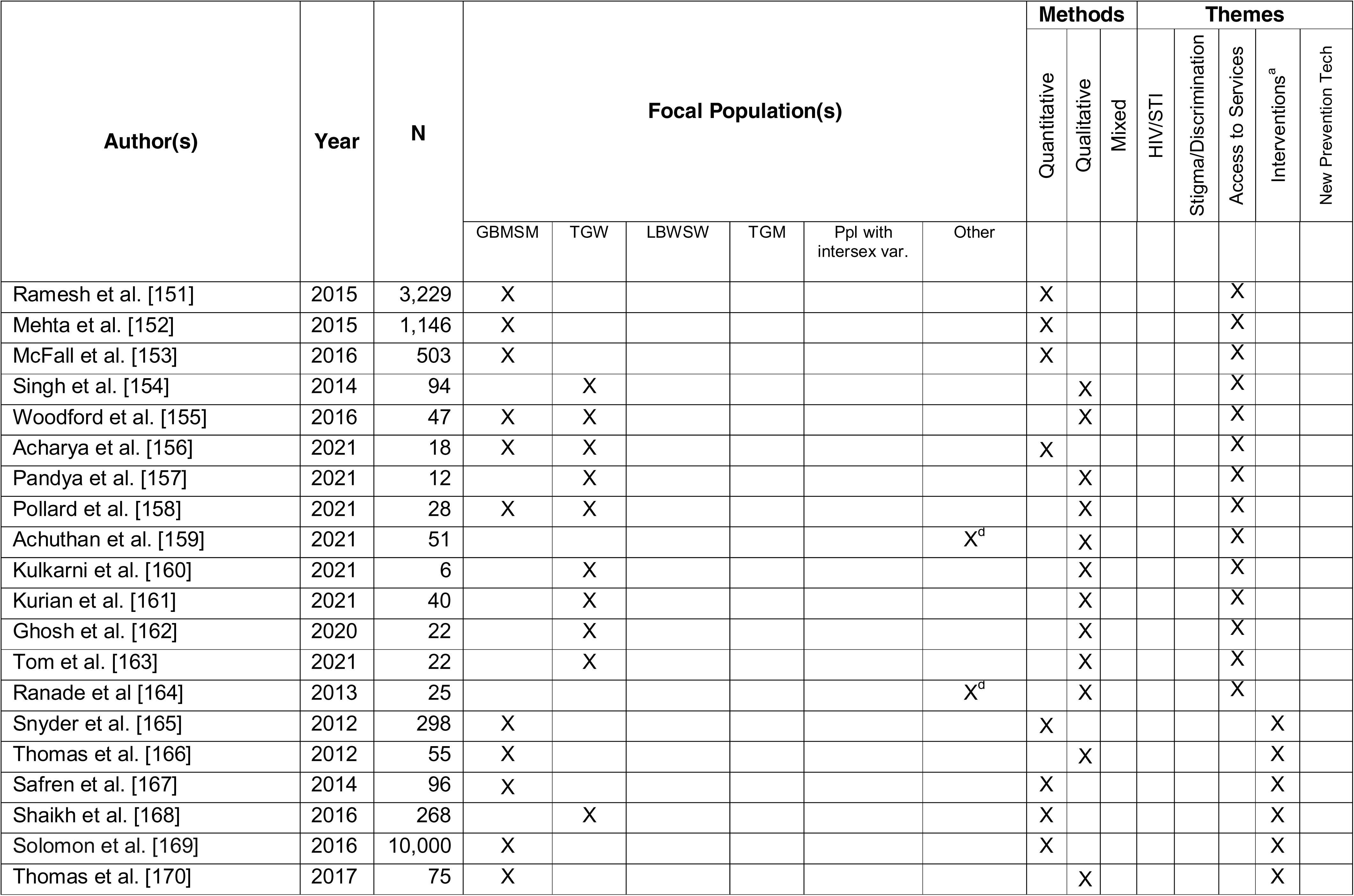
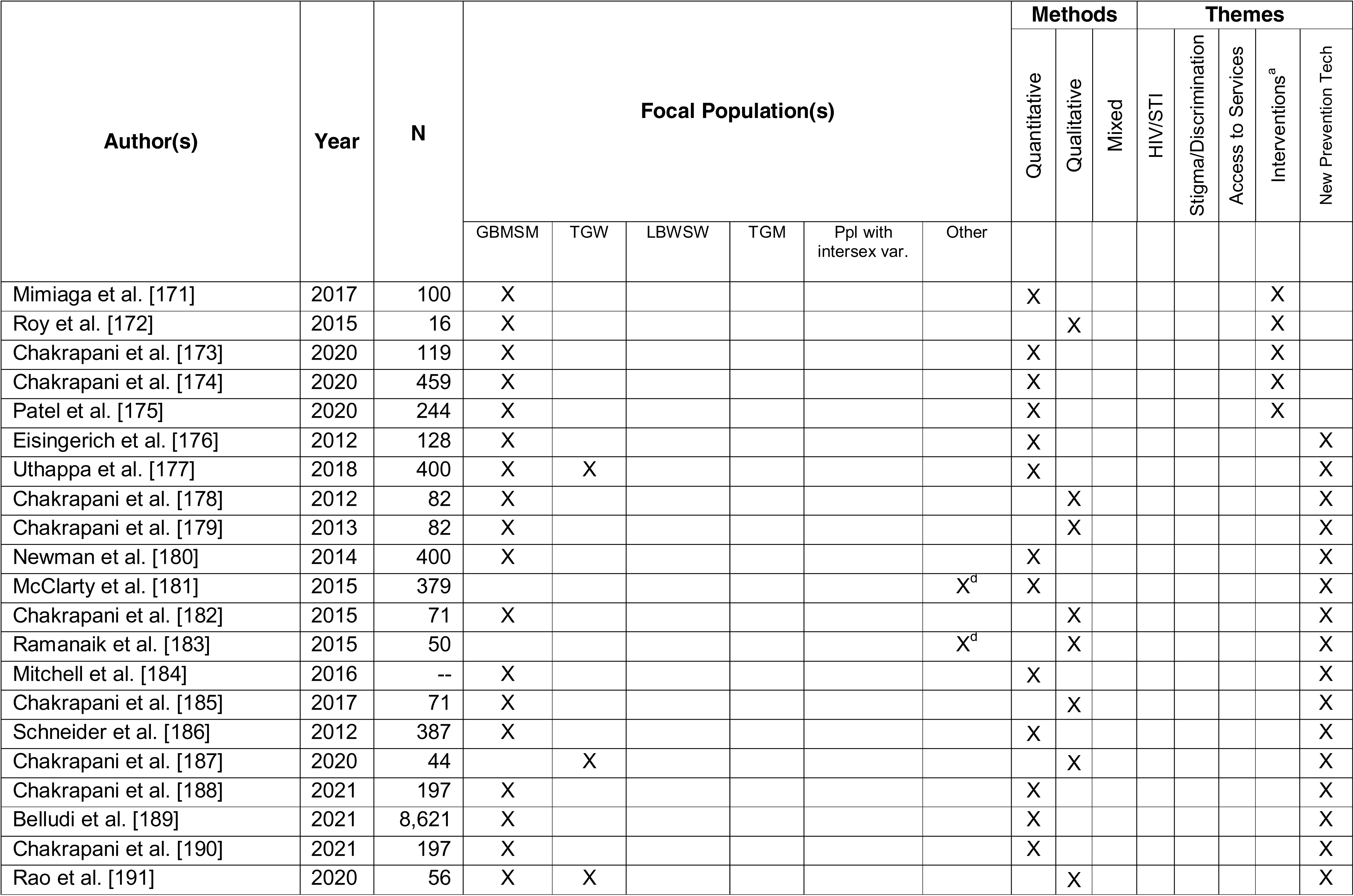
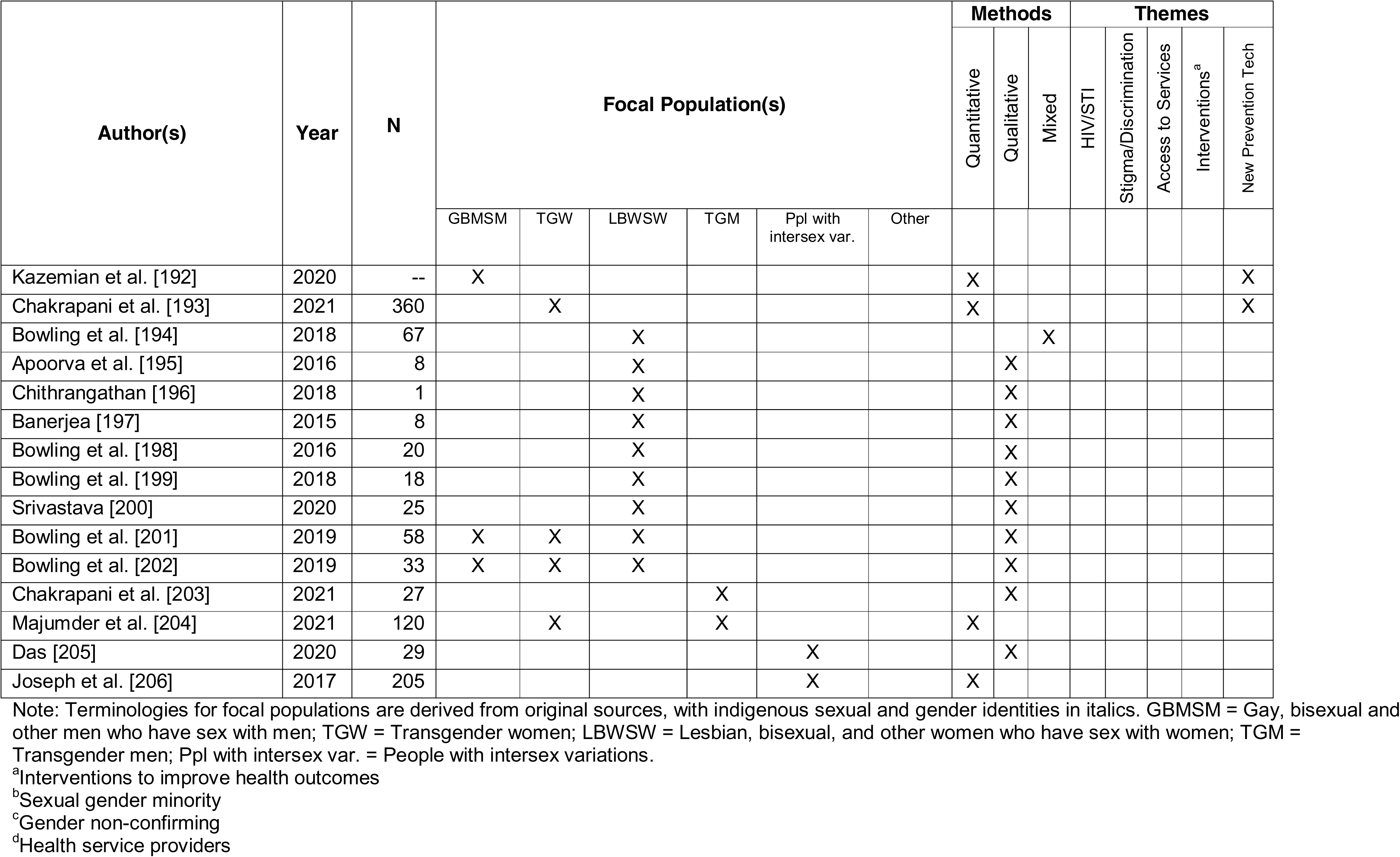
Study characteristics and themes of inclusion (n = 177)

| Author(s) | Year | N | Focal Population(s) |  |  |  |  |  | Methods |  |  | Themes |  |  |  |  |
| --- | --- | --- | --- | --- | --- | --- | --- | --- | --- | --- | --- | --- | --- | --- | --- | --- |
|  |  |  |  |  |  |  |  |  | Quantitative | Qualitative | Mixed | HIV/STI | Stigma/Discrimination | Access to Services | Interventions<sup>a</sup> | New Prevention Tech |
|  |  |  | GBMSM | TGW | LBWSW | TGM | Ppl with intersex var. | Other |  |  |  |  |  |  |  |  |
| Solomon et al. [30] | 2010 | 721 | X |  |  |  |  |  | X |  |  | X |  |  |  |  |
| Ghosh et al. [31] | 2011 | 32 | X |  |  |  |  |  | X |  |  | X |  |  |  |  |
| Sahastrabuddhe et al. [32] | 2012 | 84 |  | X |  |  |  |  | X |  |  | X |  |  |  |  |
| Ghosh et al. [33] | 2012 | 26 | X |  |  |  |  |  | X |  |  | X |  |  |  |  |
| Solomon et al. [34] | 2015 | 12,022 | X |  |  |  |  |  | X |  |  | X |  |  |  |  |
| Mayer et al. [35] | 2015 | 307 | X |  |  |  |  |  | X |  |  | X |  |  |  |  |
| Hernandez et al. [36] | 2016 | 300 | X |  |  |  |  |  | X |  |  | X |  |  |  |  |
| Aggarwal et al. [37] | 2016 | 52 | X |  |  |  |  |  | X |  |  | X |  |  |  |  |
| Solomon et al. [38] | 2016 | 12,022 | X |  |  |  |  |  | X |  |  | X |  |  |  |  |
| Raghavendran et al. [39] | 2017 | 300 | X |  |  |  |  |  | X |  |  | X |  |  |  |  |
| Hussain et al. [40] | 2018 | 277 | X |  |  |  |  |  | X |  |  | X |  |  |  |  |
| Gupte et al. [41] | 2011 | 2,633 | X | X |  |  |  |  | X |  |  | X |  |  |  |  |
| Clipman et al. [42] | 2020 | 4,994 | X |  |  |  |  |  | X |  |  | X |  |  |  |  |
| Haldar et al. [43] | 2020 | 2,584 | X |  |  |  |  |  | X |  |  | X |  |  |  |  |
| Hernandez et al. [44] | 2021 | 302 | X |  |  |  |  |  | X |  |  | X |  |  |  |  |
| Palakkal et al. [45] | 2020 | 560 | X |  |  |  |  |  | X |  |  | X |  |  |  |  |
| Prabhu et al. [46] | 2021 | 1,639 | X |  |  |  |  |  | X |  |  | X |  |  |  |  |
| Patel et al. [47] | 2021 | 20,002 | X |  |  |  |  |  | X |  |  | X |  |  |  |  |
| Kumta et al. [48] | 2010 | 831 | X |  |  |  |  |  | X |  |  | X |  |  |  |  |
| Phillips et al. [49] | 2010 | 357 | X |  |  |  |  |  | X |  |  | X |  |  |  |  |
|  |  |  |  |  |  |  |  |  | Quantitative | Qualitative | Mixed | HIV/STI | Stigma/Discrimination | Access to Services | Interventions<sup>a</sup> | New Prevention Tech |
|  |  |  | GBMSM | TGW | LBWSW | TGM | Ppl with intersex var. | Other |  |  |  |  |  |  |  |  |
| Solomon et al. [50] | 2010 | 781 | X |  |  |  |  |  |  |  | X | X |  |  |  |  |
| Setia et al. [51] | 2010 | 511 | X |  |  |  |  |  | X |  |  | X |  |  |  |  |
| Gutierrez et al. [52] | 2010 | 4,321 | X |  |  |  |  |  | X |  |  | X |  |  |  |  |
| Lorway et al. [53] | 2010 | 120 | X |  |  |  |  |  | X |  |  | X |  |  |  |  |
| Mimiaga et al. [54] | 2011 | 210 | X |  |  |  |  |  | X |  |  | X |  |  |  |  |
| Hendriksen et al. [55] | 2011 | 5 | X |  |  |  |  |  |  | X |  | X |  |  |  |  |
| Hemmige et al. [56] | 2011 | 676 | X |  |  |  |  |  | X |  |  | X |  |  |  |  |
| Lorway et al. [57] | 2011 | -- | X |  |  |  |  |  |  | X |  | X |  |  |  |  |
| Thomas et al. [58] | 2012 | 210 | X |  |  |  |  |  | X |  |  | X |  |  |  |  |
| Tomori et al. [59] | 2018 | 47 | X |  |  |  |  |  |  | X |  | X |  |  |  |  |
| Wilkerson et al. [60] | 2018 | 433 | X |  |  |  |  |  | X |  |  | X |  |  |  |  |
| Srivastava et al. [61] | 2019 | 10 | X |  |  |  |  |  |  | X |  | X |  |  |  |  |
| Mimiaga et al. [62] | 2013 | 150 | X |  |  |  |  |  | X |  |  | X |  |  |  |  |
| Saggurti et al. [63] | 2013 | 2,399 | X | X |  |  |  |  | X |  |  | X |  |  |  |  |
| Chakrapani et al. [64] | 2013 | 88 | X |  |  |  |  |  |  | X |  | X |  |  |  |  |
| Ramanathan et al. [65] | 2013 | 1618 | X |  |  |  |  |  | X |  |  | X |  |  |  |  |
| Narayanan et al. [66] | 2013 | 483 | X |  |  |  |  |  | X |  |  | X |  |  |  |  |
| Kumar et al. [67] | 2014 | 3,229 | X |  |  |  |  |  | X |  |  | X |  |  |  |  |
| Yadav et al. [68] | 2014 | 3,880 | X |  |  |  |  |  | X |  |  | X |  |  |  |  |
| Ramesh et al. [69] | 2014 | 1,608 | X |  |  |  |  |  | X |  |  | X |  |  |  |  |
|  |  |  |  |  |  |  |  |  | Quantitative | Qualitative | Mixed | HIV/STI | Stigma/Discrimination | Access to Services | Interventions<sup>a</sup> | New Prevention Tech |
|  |  |  | GBMSM | TGW | LBWSW | TGM | Ppl with intersex var. | Other |  |  |  |  |  |  |  |  |
| Mitchell et al. [70] | 2014 | 595 | X |  |  |  |  |  | X |  |  | X |  |  |  |  |
| Closson et al. [71] | 2014 | 32 | X |  |  |  |  |  |  | X |  | X |  |  |  |  |
| Ramanathan et al. [72] | 2014 | 1,305 | X | X |  |  |  |  | X |  |  | X |  |  |  |  |
| Godbole et al. [73] | 2014 | 4,682 | X |  |  |  |  |  | X |  |  | X |  |  |  |  |
| Mitchell et al. [74] | 2014 | 320 | X |  |  |  |  |  | X |  |  | X |  |  |  |  |
| Saha et al. [75] | 2015 | 227 | X |  |  |  |  |  | X |  |  | X |  |  |  |  |
| Saha et al. [76] | 2015 | 243 | X |  |  |  |  |  | X |  |  | X |  |  |  |  |
| Ramakrishnan et al. [77] | 2015 | 3,833 | X |  |  |  |  |  | X |  |  | X |  |  |  |  |
| Mahapatra et al. [78] | 2015 | 1,237 | X |  |  |  |  |  | X |  |  | X |  |  |  |  |
| Shaw et al. [79] | 2016 | 456 | X | X |  |  |  |  | X |  |  | X |  |  |  |  |
| Tomori et al. [80] | 2016 | 12,151 | X |  |  |  |  |  |  |  | X | X |  |  |  |  |
| Sinha et al. [81] | 2017 | 90 |  | X |  |  |  |  | X |  |  | X |  |  |  |  |
| Banik et al. [82] | 2019 | 72 | X |  |  |  |  |  |  | X |  | X |  |  |  |  |
| Deshpande et al. [83] | 2015 | 689 | X |  |  |  |  |  | X |  |  | X |  |  |  |  |
| Chakrapani et al. [84] | 2015 | 151 | X | X |  |  |  |  |  |  | X | X |  |  |  |  |
| Dodge et al. [85] | 2016 | 72 | X |  |  |  |  |  |  |  | X | X |  |  |  |  |
| Willie et al. [86] | 2017 | 299 |  | X |  |  |  |  | X |  |  | X |  |  |  |  |
| Ferguson et al. [87] | 2016 | 30 | X | X |  |  |  |  |  | X |  | X |  |  |  |  |
| Banik et al. [88] | 2014 | 36 | X |  |  |  |  |  |  | X |  | X |  |  |  |  |
| Wilkerson et al. [89] | 2019 | 449 | X | X |  |  |  |  | X |  |  | X |  |  |  |  |

| Author(s) | Year | N | Focal Population(s) |  |  |  |  |  | Methods |  |  | Themes |  |  |  |
| --- | --- | --- | --- | --- | --- | --- | --- | --- | --- | --- | --- | --- | --- | --- | --- |
|  |  |  |  |  |  |  |  |  | Quantitative | Qualitative | Mixed | HIV/STI | Stigma/Discrimination | Access to Services | Interventions<sup>a</sup> |
|  |  |  | GBMSM | TGW | LBWSW | TGM | Ppl with intersex var. | Other |  |  |  |  |  |  |  |
| Bhambhani et al. [90] | 2021 | 4,321 | X |  |  |  |  |  | X |  |  | X |  |  |  |
| Sudharshan et al. [91] | 2020 | 33 | X |  |  |  |  |  | X |  |  | X |  |  |  |
| Safren et al. [92] | 2021 | 608 | X |  |  |  |  |  | X |  |  | X |  |  |  |
| Kumar et al. [93] | 2020 | 23,081 | X |  |  |  |  |  | X |  |  | X |  |  |  |
| Rajan et al. [94] | 2020 | 3,325 |  | X |  |  |  |  | X |  |  | X |  |  |  |
| Sivasubramanian et al. [95] | 2011 | 150 | X |  |  |  |  |  | X |  |  |  | X |  |  |
| Logie et al. [96] | 2012 | 200 | X |  |  |  |  |  | X |  |  |  | X |  |  |
| Shaw et al. [97] | 2012 | 543 | X | X |  |  |  |  | X |  |  |  | X |  |  |
| Tomori et al. [98] | 2018 | 484 | X |  |  |  |  |  |  | X |  |  | X |  |  |
| Tomori et al. [99] | 2018 | 11,771 | X |  |  |  |  |  | X |  |  |  | X |  |  |
| Thaker et al. [100] | 2018 | 225 | X | X |  |  |  |  | X |  |  |  | X |  |  |
| Chakrapani et al. [101] | 2019 | 300 |  | X |  |  |  |  | X |  |  |  | X |  |  |
| Thompson et al. [102] | 2013 | 39 | X |  |  |  |  |  |  | X |  |  | X |  |  |
| Elouard et al. [103] | 2013 | 11 | X |  |  |  |  |  |  | X |  |  | X |  |  |
| Maroky et al. [104] | 2015 | 51 | X |  |  |  |  |  | X |  |  |  | X |  |  |
| Mimiaga et al. [105] | 2015 | 55 | X |  |  |  |  |  |  | X |  |  | X |  |  |
| Tomori et al. [106] | 2016 | 12,355 | X |  |  |  |  |  |  |  | X |  | X |  |  |
| Tomori et al. [107] | 2016 | 363 | X |  |  |  |  |  |  | X |  |  | X |  |  |
| Chakrapani et al. [108] | 2017 | 600 | X | X |  |  |  |  | X |  |  |  | X |  |  |
| Ganju et al. [109] | 2016 | 68 |  | X |  |  |  |  |  | X |  |  | X |  |  |
| Chakrapani et al. [110] | 2017 | 600 | X | X |  |  |  |  | X |  |  |  | X |  |  |
| Kalra et al. [111] | 2013 | 50 |  | X |  |  |  |  | X |  |  |  | X |  |  |
|  |  |  |  |  |  |  |  |  | Quantitative | Qualitative | Mixed | HIV/STI | Stigma/Discrimination | Access to Services | Interventions<sup>a</sup> | New Prevention Tech |
|  |  |  | GBMSM | TGW | LBWSW | T<br>G<br>M | Ppl with intersex<br>var. | Other |  |  |  |  |  |  |  |  |
| Lorway et al. [112] | 2013 | 70 | X |  |  |  |  |  |  | X |  |  | X |  |  |  |
| Manian [113] | 2014 | 10 | X | X |  |  |  |  |  | X |  |  | X |  |  |  |
| Chakrapani et al. [114] | 2017 | 300 |  | X |  |  |  |  |  | X |  |  | X |  |  |  |
| Chakrapani et al. [115] | 2018 | 40 | X |  |  |  |  |  |  | X |  |  | X |  |  |  |
| Rao et al. [116] | 2018 | 227 |  |  |  |  |  | X<sup>b</sup> | X |  |  |  | X |  |  |  |
| Pandya [117] | 2010 | 250 | X |  |  |  |  |  |  | X |  |  | X |  |  |  |
| Dutta et al. [118] | 2019 | 41 |  | X |  |  |  |  |  | X |  |  | X |  |  |  |
| Bowling et al. [119] | 2019 | 58 | X | X | X |  |  |  |  | X |  |  | X |  |  |  |
| Rao et al. [120] | 2020 | 170 |  |  |  |  |  | X<sup>b</sup> | X |  |  |  | X |  |  |  |
| Li et al. [121] | 2017 | 487 | X | X |  |  |  |  |  |  | X |  | X |  |  |  |
| Chavada et al. [122] | 2021 | 100 |  | X |  |  |  |  | X |  |  |  | X |  |  |  |
| Bhattacharya et al. [123] | 2020 | 98 | X | X |  |  |  |  | X |  |  |  | X |  |  |  |
| Dhabhar et al. [124] | 2020 | 112 | X |  | X |  |  |  | X |  |  |  | X |  |  |  |
| Banerjee et al. [125] | 2020 | 10 |  | X |  |  |  |  |  | X |  |  | X |  |  |  |
| Azhar et al. [126] | 2021 | 16 |  |  |  |  |  | X<sup>c</sup> |  |  | X |  | X |  |  |  |
| Srivastava et al. [127] | 2021 | 20 |  | X |  |  |  |  |  | X |  |  | X |  |  |  |
| Prabhu et al. [128] | 2020 | 1,454 | X |  |  |  |  |  | X |  |  |  | X |  |  |  |
| Arvind et al. [129] | 2021 | 20 |  | X |  |  |  |  |  |  | X |  | X |  |  |  |
| Thirunavukkarasu et al. [130] | 2021 | 235 | X |  |  |  |  |  | X |  |  |  | X |  |  |  |
| Sharma et al. [131] | 2020 | 296 | X | X | X |  |  |  |  |  | X |  | X |  |  |  |
|  |  |  |  |  |  |  |  |  | Quantitative | Qualitative | Mixed | HIV/STI | Stigma/Discrimination | Access to Services | Interventions<sup>a</sup> | New Prevention Tech |
|  |  |  | GBMSM | TGW | LBWSW | TGM | Ppl with intersex var. | Other |  |  |  |  |  |  |  |  |
| Dhaor [132] | 2021 | 21 | X | X |  |  |  |  |  |  | X |  | X |  |  |  |
| Safren et al. [133] | 2021 | 608 | X |  |  |  |  |  | X |  |  |  | X |  |  |  |
| Pufahl et al. [134] | 2021 | 184 |  |  |  |  |  | X<sup>c</sup> | X |  |  |  | X |  |  |  |
| Majumder et al. [135] | 2020 | 37 |  | X |  |  |  |  | X |  |  |  | X |  |  |  |
| Joshi et al. [136] | 2021 | 33 |  | X |  |  |  |  | X |  |  |  | X |  |  |  |
| Jesus et al. [137] | 2020 | 23 |  | X |  |  |  |  |  |  | X |  | X |  |  |  |
| Sharma et al. [138] | 2020 | 207 | X |  |  |  |  |  |  |  | X |  | X |  |  |  |
| Sartaj et al. [139] | 2021 | 50 |  | X |  |  |  |  | X |  |  |  | X |  |  |  |
| Srivastava et al. [140] | 2021 | 3,548 | X | X |  |  |  |  | X |  |  |  | X |  |  |  |
| Jethwani et al. [141] | 2014 | 124 | X |  |  |  |  |  | X |  |  |  | X |  |  |  |
| Singh et al. [142] | 2018 | 15 | X |  |  |  |  |  | X |  |  |  | X |  |  |  |
| Mogasale et al. [143] | 2010 | -- | X | X |  |  |  |  | X |  |  |  |  | X |  |  |
| Chakrapani et al. [144] | 2011 | 38 | X | X |  |  |  |  |  | X |  |  |  | X |  |  |
| Gurung et al. [145] | 2011 | 8,9621 | X | X |  |  |  |  | X |  |  |  |  | X |  |  |
| Woodford et al. [146] | 2012 | 132 | X |  |  |  |  |  | X |  |  |  |  | X |  |  |
| Beattie et al. [147] | 2012 | 90 | X | X |  |  |  |  |  | X |  |  |  | X |  |  |
| Pina et al. [148] | 2018 | 300 | X | X |  |  |  |  | X |  |  |  |  | X |  |  |
| Patel et al. [149] | 2018 | 4,179 | X |  |  |  |  |  | X |  |  |  |  | X |  |  |
| Samuel et al. [150] | 2018 | 212 |  | X |  |  |  |  | X |  |  |  |  | X |  |  |
|  |  |  |  |  |  |  |  |  | Quantitative | Qualitative | Mixed | HIV/STI | Stigma/Discrimination | Access to Services | Interventions<sup>a</sup> | New Prevention Tech |
|  |  |  | GBMSM | TGW | LBWSW | TGM | Ppl with intersex var. | Other |  |  |  |  |  |  |  |  |
| Ramesh et al. [151] | 2015 | 3,229 | X |  |  |  |  |  | X |  |  |  |  | X |  |  |
| Mehta et al. [152] | 2015 | 1,146 | X |  |  |  |  |  | X |  |  |  |  | X |  |  |
| McFall et al. [153] | 2016 | 503 | X |  |  |  |  |  | X |  |  |  |  | X |  |  |
| Singh et al. [154] | 2014 | 94 |  | X |  |  |  |  |  | X |  |  |  | X |  |  |
| Woodford et al. [155] | 2016 | 47 | X | X |  |  |  |  |  | X |  |  |  | X |  |  |
| Acharya et al. [156] | 2021 | 18 | X | X |  |  |  |  | X |  |  |  |  | X |  |  |
| Pandya et al. [157] | 2021 | 12 |  | X |  |  |  |  |  | X |  |  |  | X |  |  |
| Pollard et al. [158] | 2021 | 28 | X | X |  |  |  |  |  | X |  |  |  | X |  |  |
| Achuthan et al. [159] | 2021 | 51 |  |  |  |  |  | X<sup>d</sup> |  | X |  |  |  | X |  |  |
| Kulkarni et al. [160] | 2021 | 6 |  | X |  |  |  |  |  | X |  |  |  | X |  |  |
| Kurian et al. [161] | 2021 | 40 |  | X |  |  |  |  |  | X |  |  |  | X |  |  |
| Ghosh et al. [162] | 2020 | 22 |  | X |  |  |  |  |  | X |  |  |  | X |  |  |
| Tom et al. [163] | 2021 | 22 |  | X |  |  |  |  |  | X |  |  |  | X |  |  |
| Ranade et al [164] | 2013 | 25 |  |  |  |  |  | X<sup>d</sup> |  | X |  |  |  | X |  |  |
| Snyder et al. [165] | 2012 | 298 | X |  |  |  |  |  | X |  |  |  |  |  | X |  |
| Thomas et al. [166] | 2012 | 55 | X |  |  |  |  |  |  | X |  |  |  |  | X |  |
| Safren et al. [167] | 2014 | 96 | X |  |  |  |  |  | X |  |  |  |  |  | X |  |
| Shaikh et al. [168] | 2016 | 268 |  | X |  |  |  |  | X |  |  |  |  |  | X |  |
| Solomon et al. [169] | 2016 | 10,000 | X |  |  |  |  |  | X |  |  |  |  |  | X |  |
| Thomas et al. [170] | 2017 | 75 | X |  |  |  |  |  |  | X |  |  |  |  | X |  |
|  |  |  |  |  |  |  |  |  | Quantitative | Qualitative | Mixed | HIV/STI | Stigma/Discrimination | Access to Services | Interventions<sup>a</sup> | New Prevention Tech |
|  |  |  | GBMSM | TGW | LBWSW | TGM | Ppl with intersex var. | Other |  |  |  |  |  |  |  |  |
| Mimiaga et al. [171] | 2017 | 100 | X |  |  |  |  |  | X |  |  |  |  |  | X |  |
| Roy et al. [172] | 2015 | 16 | X |  |  |  |  |  |  | X |  |  |  |  | X |  |
| Chakrapani et al. [173] | 2020 | 119 | X |  |  |  |  |  | X |  |  |  |  |  | X |  |
| Chakrapani et al. [174] | 2020 | 459 | X |  |  |  |  |  | X |  |  |  |  |  | X |  |
| Patel et al. [175] | 2020 | 244 | X |  |  |  |  |  | X |  |  |  |  |  | X |  |
| Eisingerich et al. [176] | 2012 | 128 | X |  |  |  |  |  | X |  |  |  |  |  |  | X |
| Uthappa et al. [177] | 2018 | 400 | X | X |  |  |  |  | X |  |  |  |  |  |  | X |
| Chakrapani et al. [178] | 2012 | 82 | X |  |  |  |  |  |  | X |  |  |  |  |  | X |
| Chakrapani et al. [179] | 2013 | 82 | X |  |  |  |  |  |  | X |  |  |  |  |  | X |
| Newman et al. [180] | 2014 | 400 | X |  |  |  |  |  | X |  |  |  |  |  |  | X |
| McClarty et al. [181] | 2015 | 379 |  |  |  |  |  | X<sup>d</sup> | X |  |  |  |  |  |  | X |
| Chakrapani et al. [182] | 2015 | 71 | X |  |  |  |  |  |  | X |  |  |  |  |  | X |
| Ramanaik et al. [183] | 2015 | 50 |  |  |  |  |  | X<sup>d</sup> |  | X |  |  |  |  |  | X |
| Mitchell et al. [184] | 2016 | -- | X |  |  |  |  |  | X |  |  |  |  |  |  | X |
| Chakrapani et al. [185] | 2017 | 71 | X |  |  |  |  |  |  | X |  |  |  |  |  | X |
| Schneider et al. [186] | 2012 | 387 | X |  |  |  |  |  | X |  |  |  |  |  |  | X |
| Chakrapani et al. [187] | 2020 | 44 |  | X |  |  |  |  |  | X |  |  |  |  |  | X |
| Chakrapani et al. [188] | 2021 | 197 | X |  |  |  |  |  | X |  |  |  |  |  |  | X |
| Belludi et al. [189] | 2021 | 8,621 | X |  |  |  |  |  | X |  |  |  |  |  |  | X |
| Chakrapani et al. [190] | 2021 | 197 | X |  |  |  |  |  | X |  |  |  |  |  |  | X |
| Rao et al. [191] | 2020 | 56 | X | X |  |  |  |  |  | X |  |  |  |  |  | X |
|  |  |  |  |  |  |  |  |  | Quantitative | Qualitative | Mixed | HIV/STI | Stigma/Discrimination | Access to Services | Interventions<sup>a</sup> | New Prevention Tech |
|  |  |  | GBMSM | TGW | LBWSW | TGM | Ppl with intersex var. | Other |  |  |  |  |  |  |  |  |
| Kazemian et al. [192] | 2020 | -- | X |  |  |  |  |  | X |  |  |  |  |  |  | X |
| Chakrapani et al. [193] | 2021 | 360 |  | X |  |  |  |  | X |  |  |  |  |  |  | X |
| Bowling et al. [194] | 2018 | 67 |  |  | X |  |  |  |  |  | X |  |  |  |  |  |
| Apoorva et al. [195] | 2016 | 8 |  |  | X |  |  |  |  | X |  |  |  |  |  |  |
| Chithrangathan [196] | 2018 | 1 |  |  | X |  |  |  |  | X |  |  |  |  |  |  |
| Banerjea [197] | 2015 | 8 |  |  | X |  |  |  |  | X |  |  |  |  |  |  |
| Bowling et al. [198] | 2016 | 20 |  |  | X |  |  |  |  | X |  |  |  |  |  |  |
| Bowling et al. [199] | 2018 | 18 |  |  | X |  |  |  |  | X |  |  |  |  |  |  |
| Srivastava [200] | 2020 | 25 |  |  | X |  |  |  |  | X |  |  |  |  |  |  |
| Bowling et al. [201] | 2019 | 58 | X | X | X |  |  |  |  | X |  |  |  |  |  |  |
| Bowling et al. [202] | 2019 | 33 | X | X | X |  |  |  |  | X |  |  |  |  |  |  |
| Chakrapani et al. [203] | 2021 | 27 |  |  |  | X |  |  |  | X |  |  |  |  |  |  |
| Majumder et al. [204] | 2021 | 120 |  | X |  | X |  |  | X |  |  |  |  |  |  |  |
| Das [205] | 2020 | 29 |  |  |  |  | X |  |  | X |  |  |  |  |  |  |
| Joseph et al. [206] | 2017 | 205 |  |  |  |  | X |  | X |  |  |  |  |  |  |  |

Note: Terminologies for focal populations are derived from original sources, with indigenous sexual and gender identities in italics. GBMSM = Gay, bisexual and other men who have sex with men; TGW = Transgender women; LBWSW = Lesbian, bisexual, and other women who have sex with women; TGM = Transgender men; Ppl with intersex var. = People with intersex variations.
<sup>a</sup>Interventions to improve health outcomes
<sup>b</sup>Sexual gender minority
<sup>c</sup>Gender non-confirming
<sup>d</sup>Health service providers

### Study characteristics

Figure 2 shows the frequency of publications by year. Of the 177 articles, the majority (59%; n = 105) were published from 2016 onward. In terms of methodology, 62% were quantitative, 31% qualitative, and 7% mixed methods studies. As shown in Figure 3, the majority (55%; n = 98) of studies were conducted among MSM, 16% (n = 28) among TGW, and 14% (n = 25) among both MSM and TGW. Seven studies (4%) were conducted among lesbian or bisexual women, five (3%) among LGBTQI+ people as a whole, and two each among transmasculine people, and people with intersex variations.

**Fig 2.**
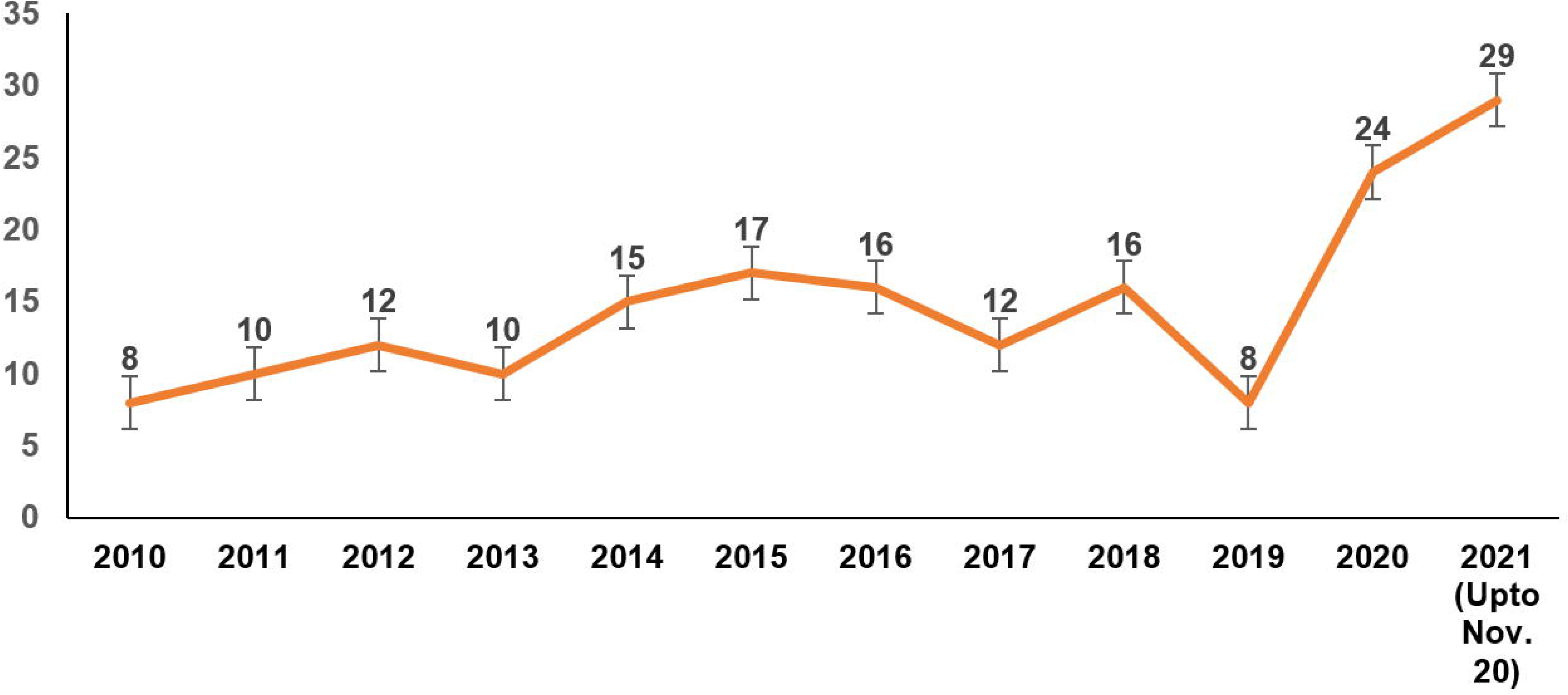
Distribution of peer-reviewed articles by year of publication (N = 177)

**Fig 3.**
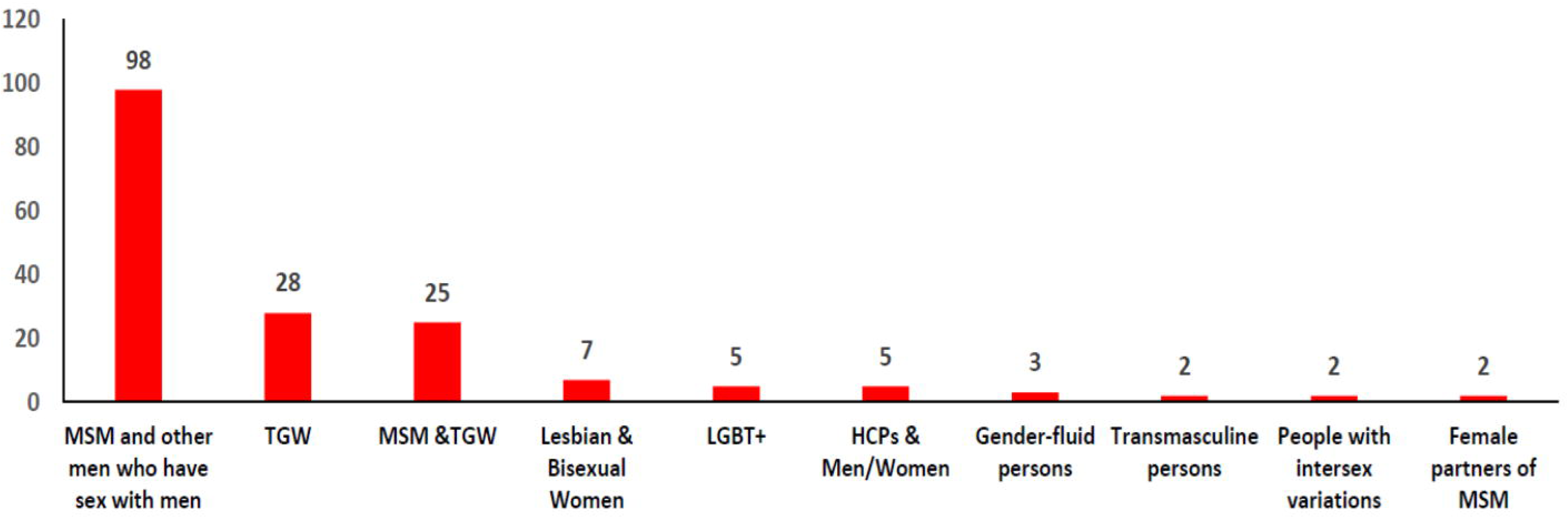
Distribution of focal populations in the peer-reviewed articles (N = 177)

Nearly half (47%; n = 84) of the studies were conducted in four (of 28) Indian states— Maharashtra (n = 30), Tamil Nadu (n = 23), Karnataka (n = 19) or Andhra Pradesh (n = 12), with the majority in these state’s capitals—Mumbai, Chennai, Bangalore, or Hyderabad. Over a third (36%; n = 65) of the studies were conducted in multiple Indian states.

### HIV/STI prevalence and risk factors

Figure 4 shows that 37% (n = 65) of the articles focused on reporting STI/HIV prevalence estimates [30–47] and correlates of HIV-related risk behaviors [48–94] among MSM and TGW. In the 18 studies [30–47] that reported HIV and STI prevalence estimates among MSM and TGW, nine [31-33, 37, 39–42, 45] were conducted in clinical settings, six [30, 34, 35, 38, 43, 46] in community settings, and three [36, 44, 47] in both clinical and community settings. Of these 18 studies, eight [30, 34, 35, 37, 40, 43, 45, 46] reported HIV/STI prevalence and risk factors among MSM, three [36, 39, 44] human papillomavirus (HPV) prevalence among MSM living with HIV, and three [31, 33, 45] reported prevalence of perianal dermatoses, HPV and other STIs (such as syphilis, chlamydia and gonorrhea) among MSM. Two studies [38, 47] reported correlates of HIV incidence among MSM, with one study each reporting Hepatitis C prevalence among MSM living with HIV [42], and one study the prevalence of herpes [45].

**Fig 4.**
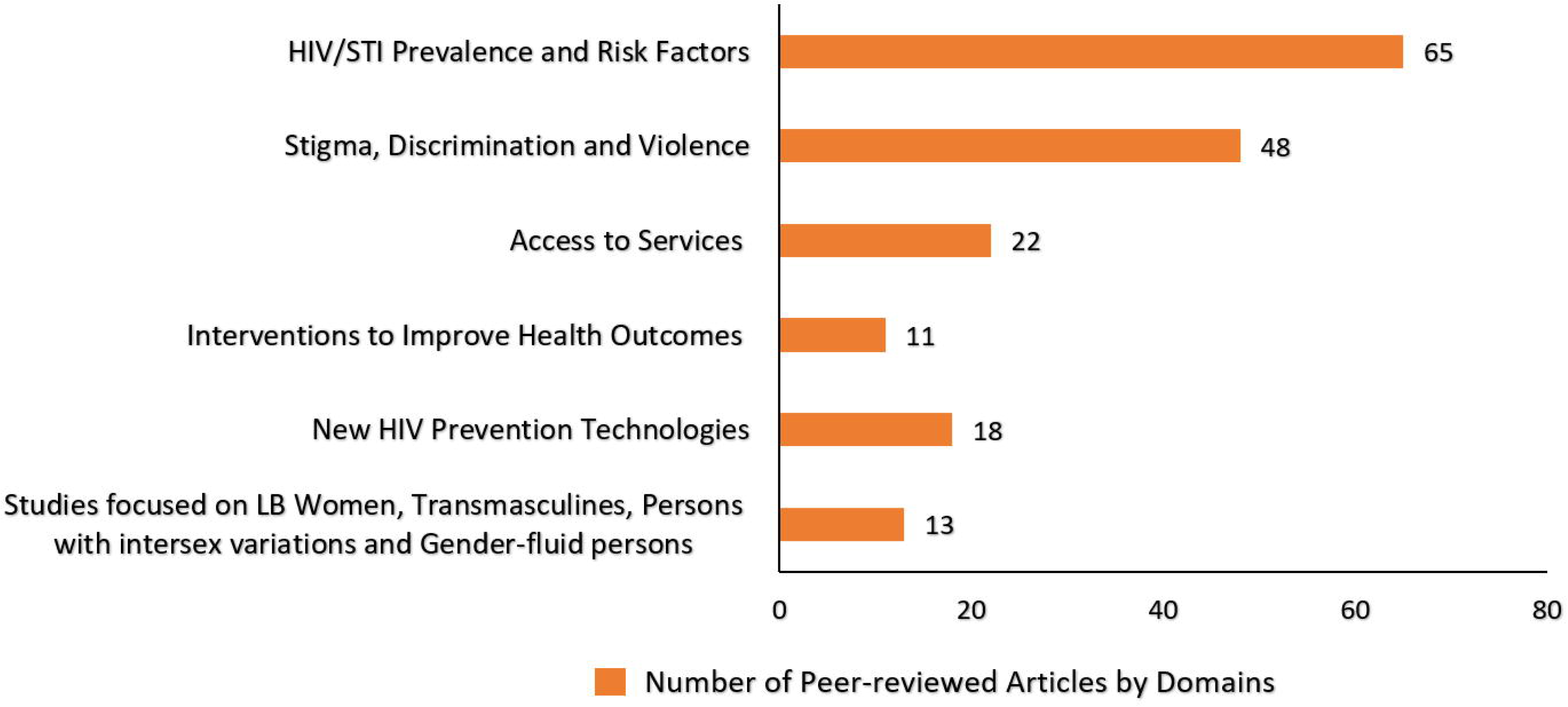
Distribution of peer-reviewed articles by domains (N=177)

Overall, HIV prevalence among MSM ranged from 3.8% to 23.0% across different study sites. Among MSM, HPV/genital warts (23.0% to 95.0%), syphilis (0.8% to 11.9%), HSV/genital herpes (7.1 to 32.0%), and genital molluscum contagium (9.6%) were the most commonly reported STIs [30, 31, 33, 34, 36, 37, 39, 40, 42, 45]. One study [42] reported Hepatitis-C prevalence among MSM as 1.3%. Syphilis rates tended to be higher among single MSM (8.3%) than married MSM (1.0%) [35]. In a study [32] conducted among 84 TGW who attended STI clinics in Pune, HIV prevalence was 45.2%.

Forty-seven articles [48–94] reported correlates of HIV-related risk among MSM and TGW. Among MSM, significant correlates of HIV risk behaviors/indicators—i.e., condomless sex [48-50, 58, 64, 77, 78, 92, 93], infrequent HIV testing [60, 65, 72, 74], and HIV/STI positivity [48, 51, 55, 79]—were low literacy and unemployment [48, 76, 77], alcohol and/or drug use [54, 60, 64, 65, 79, 90, 93], engagement in sex work [49, 60, 61, 65, 67, 68, 75, 76, 78], higher number of male sexual partners [48-50, 53, 56, 72, 74], early age of sexual debut [93], and low HIV risk perception [60, 65, 72, 74]. Six of the 47 articles included data on TGW; five of these [79, 81, 84, 87, 89] did not provide details on correlates of HIV risk behaviors, with one study [94] reporting that having a male regular partner was associated with HIV seropositivity.

### Stigma, discrimination, and violence, and health impacts

Over one-fourth of the articles (27%; n=48) [95–142] reported on stigma, discrimination, violence, and their associations with physical and mental health. Among these, 16 articles focused on stigma-related aspects of LGBTQI+ health [96, 100–102, 109, 110, 112, 114, 119, 124–126, 129, 132, 137, 140], 3 on violence [97, 103, 118], 17 on mental health and its correlates, such as quality of life [95, 99, 105, 106, 108, 111, 115, 123, 127, 128, 130, 131, 135, 136, 138, 139, 142], two on resilience [122, 133] and one article each on coping skills [141] and promoting LGBTQI+ acceptance [134]. Three articles reported on stress [116], perceived psychological impact [120] and violence [121] associated with Section-377 of the Indian Penal Code, which until September 2018 criminalised adult consensual same-sex relationships.

Several studies highlighted various types of stigma and discrimination experienced by MSM and TGW, which include perceived stigma, felt normative stigma, HIV-related stigma, family- enacted stigma, gender non-conformity stigma, and internalized stigma [96, 100, 101, 109, 124–126, 129, 132, 138], gender discrimination, workplace discrimination [137, 139] and polyvictimization [140]. Perpetrators of discrimination and violence against MSM and TGW, including those living with HIV, included peers, sexual partners, family members, healthcare providers, and police [98, 102, 103, 109, 112, 118, 119, 129, 130, 137, 139]. Fear of discrimination and suboptimal care [112] or refusal of care [109] prevented some persons from disclosing their sexual or gender identity to healthcare providers.

Fifteen studies [98, 99, 107–109, 112, 115, 125, 127–130, 137, 139, 140] indicated that stigma and discrimination contribute to depression and other negative mental health outcomes, such as suicidal ideation or attempts, among sexual and gender minorities. Two studies documented a high prevalence of mental health issues among MSM: depression (29% to 45%), anxiety (24% to 40%), suicidal ideation (45% to 53%), suicide attempts (23%), substance abuse (28%) including alcohol dependence (15% to 22%) [95, 130]. Similarly, among TGW, high levels of depression (43%), problematic alcohol use (37%) [108], anxiety (39%), depression (21%), suicide risk (75.8%) [136] and violence (52%) [139] were reported. Three studies with MSM [99, 108, 109, 115] and one with MSM and TGW [108] reported psychosocial syndemics, that is, co-occurring psychosocial conditions such as problematic alcohol use and internalized homonegativity, and their synergistic impact on HIV risk. The COVID-19 pandemic was also addressed as exacerbating psychological distress among LGBTQI+ people [125, 131].

Several studies addressed resilience, coping, and social support. A few studies documented various types of social support and other resilience resources available to MSM and TGW [107, 109, 117], with one study reporting moderate or high levels of resilience among 72% of TGW [122]. In terms of coping with adversity, MSM and TGW reported supportive roles of peers, NGOs [109], family, friends and partners [107], and gharanas (‘clans’ or houses of hijra- identified trans people) [127]. Some MSM and TGW reported strategies to prevent violence, discrimination and psychological distress, which included bribing police, running away from unsafe places and persons, and negotiating condom use during forced sex encounters [109], hiding sexual identities [103], denial [123], and behavioral disengagement [141]. One study documented positive coping strategies among older transgender people, such as spirituality, hope, and acceptance of gender dissonance [125]. In a few studies, social support and resilient coping strategies were identified as predictors of HIV risk [108] or mediators and moderators of the effects of discrimination on HIV risk or depression [110]. A resilience-based psychosocial intervention that integrated counselling was found to be effective in reducing HIV risk among MSM, with self-esteem and depressive symptoms mediating this effect [133]. A community-based theatre intervention was identified as effective in improving positive attitudes and knowledge, and promoting acceptance and solidarity towards LGBTQI+ communities among young adult heterosexual audiences [134].

### Access to services: HIV/STIs and gender-affirmative procedures

In total, 22 studies [143–164]—10 quantitative [143, 145, 146, 148–153, 156] and 12 qualitative [144, 147, 154, 155, 157–164]—investigated access to HIV/STI services, gender transition services, and other clinical services. Four of these studies focused on HIV testing [145, 148, 150, 154] and four [144, 148, 156, 162] on antiretroviral treatment (ART) access and uptake among MSM and TGW living with HIV. Two studies [152, 153] addressed the HIV care continuum and linkages to care, three [147, 157, 158] challenges in accessing HIV testing, treatment and care services among MSM and TGW. Five studies focused on access to healthcare and support services for TGW: access to gender transition services [154], barriers to dental [150] and eye care [160], gender-affirmative technologies [159], and welfare schemes for TGW [161].

In relation to HIV testing knowledge and access among MSM, quantitative studies [146, 149, 151] reported that a vast majority of those recruited through CBOs were aware of HIV testing facilities (86%) or tested for HIV (71%) [146] compared to MSM recruited through online social networking sites [149]. Factors such as high literacy levels, being 25 to 34 years, engagement in sex work and exposure to HIV intervention programs were associated with higher rates of HIV testing. Qualitative studies [147, 155] on HIV testing among MSM and TGW in two cities highlighted barriers such as HIV stigma and discrimination in healthcare settings and fears of adverse social consequences of testing HIV positive, and facilitators such as access to outreach programs operated by CBOs/NGOs, and accurate HIV risk perception.

Four studies (2 qualitative [144, 162] and 2 quantitative [148, 156]) conducted among MSM and TGW living with HIV reported that multilevel barriers prevented or significantly delayed access to free ART: the qualitative studies reported support from healthcare providers and peers as facilitators of ART adherence, while the quantitative studies [148, 156] indicated that 76% (n=65/85) were on ART and 48% of these (n=31/65) reported nonadherence [148]. Those who were younger and who had negative beliefs about ART were less likely to be adherent [148]. Low levels of knowledge, negative perceptions about ART, and ART nonadherence were significantly associated with lower levels of viral suppression [156].

In relation to access to gender transition services, a qualitative study [154] reported a near-total absence of gender-affirmative hormone therapy and gender-affirmative surgeries in public hospitals. Among three qualitative studies on challenges in accessing HIV testing and treatment services among MSM and TGW, two [157, 158] reported challenges faced by MSM and TGW in accessing HIV and gender transition-related services in the time of COVID-19.

### Interventions to improve health outcomes among LGBTQI+ populations

Eleven articles [165–175] focused on health-related interventions, especially in relation to HIV prevention, of which 10 were exclusively conducted with MSM. Six of the 12 studies were pilot studies, including four pilot RCTs [167, 171, 173, 175]. Two articles reported qualitative formative research studies to design counselling-based [166] and mobile phone-based interventions [170]. Studies of interventions to increase condom use or HIV testing utilized diverse modalities, such as face-to-face risk reduction counseling [167], provision of community-friendly services [168], virtual counseling [165], internet-based [175] and mobile phone-based messages [171], and motivational interviewing techniques [173, 174]. Other intervention studies used video-based technologies such as mobile game-based learning for peer education [172], and a video-based counseling session [165].

### New HIV prevention technologies and their acceptability

Overall, 18 studies [176–193] (11 quantitative, 7 qualitative) focused on new HIV prevention technologies, including oral pre-exposure prophylaxis (PrEP) [176, 177, 182, 184, 187–190, 192, 193], future HIV vaccines [178–181, 183] and rectal microbicides [185], as well as medical male circumcision [186], and oral HIV self-testing [191].

Of the ten articles on PrEP, eight examined acceptability or willingness to use PrEP among MSM and TGW; one explored the impact of prioritizing PrEP for MSM [184], and one compared the cost-effectiveness of offering PrEP to MSM with semiannual HIV testing as opposed to WHO-recommended 3-month testing [192]. Quantitative studies [176, 177, 188–190, 193] reported generally high willingness to use PrEP among MSM and TGW despite low levels of awareness. Qualitative studies [183, 188] reported correlates of PrEP uptake, including perceived effectiveness in serodiscordant relationships, offering protection in cases of forced sex encounters, ability to use covertly, ability to have sex without condoms and anxiety-less sex; barriers included PrEP stigma, fear of disclosure to one’s family or partners/spouse, and being labelled as HIV-positive or ‘promiscuous’ by peers. A mathematical modelling study [184] in Bangalore reported that PrEP could prevent a substantial proportion of infections among MSM (27% of infections over 10 years, with 60% coverage and 50% adherence).

Of the 5 studies on future HIV vaccine acceptability [178–181, 183], two [178, 180] assessed willingness to participate (WTP) in hypothetical HIV vaccine trials among MSM; one [179] explored mental models of candidate HIV vaccines and clinical trials; and two [181, 183] assessed frontline health service providers’ perspectives on HIV vaccine trials and their likelihood of recommending HIV vaccines to MSM populations.

### Underrepresented LGBTQI+ populations: Sexual minority women, transmasculine people and people with intersex variations

#### Sexual minority women

Seven studies (4%) focused on sexual minority women [194–200], while two additional studies [201, 202] included sexual minority women as part of a larger sample. Among the seven studies, most focused on romantic relationships, such as communication and prioritization in relationships [199], difficulties in maintaining relationships [196], understanding in intimate relationships [197, 198] and lack of legal recognition of same-gender romantic partnerships [198]. One study [197] used a collaborative ethnographic approach to capture the understanding of community and activism from the perspectives of “women loving women” which had indirect connections to mental health. Another study [200] documented resilience sources (e.g., self-confidence, optimism) used by sexual minority women to cope with major stressors.

The sexual health of sexual minority women was explored in two studies [194, 198]. One used photo-elicitation interviews and a survey to explore health behaviors and concerns [194], reporting that a vast majority of sexual minority women were not accessing preventive healthcare services such as breast or cervical cancer screening, and most had never been tested for STIs. The other study [198] reported lack of knowledge regarding STIs and difficulty in identifying LGBTQ-friendly service providers as major barriers to accessing preventive services.

#### Transmasculine people

Two studies (1%) [203, 204] focused on transmasculine people’s health: one [203] documented challenges in negotiating gender identity in various spaces, such as family, educational settings, workplace and neighborhoods; and one [204] reported that a substantially higher proportion of transmasculine persons (36.3%) attempted suicide when compared with transfeminine persons (24.7%).

#### People with intersex variations

Among the two studies (1%) [205, 206] that focused on people with intersex variations, one [205] examined how healthcare professionals decide on gender assignment of intersex children, and the other study [206] documented the social stigma faced by people with intersex variations and their families. Findings from both of these studies highlighted that gender assignment decisions are influenced by sociocultural factors: parents of intersex children preferred a male gender assignment possibly because of the social advantages of growing up as a male in a patriarchal society.

## DISCUSSION

This scoping review of a decade of peer-reviewed research on the health of LGBTQI+ people in India demonstrates a trend of increased publications addressing the health of sexual and gender minorities; however, it also identifies substantial gaps in the research—in terms of focal populations, geographical coverage, health conditions, and methods. Overall, this review demonstrates a predominant focus on HIV and HIV-related risk behaviors among MSM and TGW populations; of these, a small subset were intervention studies aiming to improve the health of MSM and TGW. Notably, this review reveals the near complete omission of research on the health of sexual minority women—less than 4% of the studies identified. And amid the substantial focus on transgender women, largely in the context of HIV, scant research addressed the health of transmasculine people.

From a methodological perspective, among the quantitative studies that constituted the majority of the research, most were cross-sectional and descriptive in nature; few studies used longitudinal designs or mixed methods approaches, with very few intervention trials. The inclusion of a substantial proportion of qualitative and mixed methods studies, however, suggests a strength in the potential for characterizing the lived experiences of diverse LGBTQI+ people and experiences in the context of health disparities and challenges in healthcare access. Nevertheless, these too were dominated by a focus on MSM and TGW.

The persistent and substantial gaps identified, even amid the overall increase in LGBTQI+ health research in India, have important implications for future research and research funding, health policies and programs, and healthcare services and practices for LGBTQI+ populations. There is a clear need to expand the evidence base on LGBTQI+ health in India to the many health and mental health conditions beyond HIV, and to the health challenges experienced across the diversity of LGBTQI+ people.

Specific population gaps identified in health research among LGBTQI+ people in India indicate the need for greater attention to lesbian and bisexual women, including potential health and mental health disparities compared to cisgender heterosexual women, amid the vastly disproportionate focus on the health of sexual minority men. Additional focus on lesbian and bisexual women’s experiences in access to and use of health services is sorely needed across an array of health conditions and healthcare settings, particularly given studies that reported underutilization of routine preventive healthcare services. Further gaps emerged in the dearth of research with transmasculine people [207], and more broadly in research on access to medical and surgical gender-affirmative care needs for trans people. Greater attention to studies of healthcare providers and healthcare settings that aim to improve access to gender-affirmative clinical services are needed. Finally, there is a wholesale lack of health research among people with intersex variations. Future studies should focus on general health profiles, experiences in access to healthcare, and impact of non-essential or ‘corrective’ surgeries on health and mental health outcomes [208, 209].

Overall, the relatively small number of intervention studies were largely conducted with MSM in relation to HIV prevention. Nevertheless, while NACO supports several targeted interventions among MSM and TGW, with estimated programmatic coverage of nearly 88% to 95% of at-risk MSM and TGW [210], the lack of peer-reviewed publications on the effectiveness of such interventions limits their contribution to evidence-informed HIV prevention programmes and policies in India. Although these interventions are primarily for programmatic purposes, the absence of published data represents a missed opportunity.

The stark lack of formal health outreach structures in India for lesbian and bisexual women, and for transmasculine people, makes it challenging to reach these populations through established organizational partners. Accordingly, greater involvement of a diversity of LGBTQI+ community-led groups in collaborative and participatory research studies is needed to expand opportunities to engage their inputs on research priorities, recruitment, and data collection methods, thereby also building their capacity in guiding and implementing research. Such participatory mechanisms may be key to meaningful involvement of diverse and under- represented groups among LGBTQI+ communities and expanding relevant research evidence to advance their health. Strategic research funding mechanisms that target such underrepresented groups as well as requiring community partnerships in certain health research streams may be mechanisms to support such initiatives moving forward.

This synthesis also highlights the connections between stigma, discrimination and violence, and the health issues faced by LGBTQI+ people. Several studies advance evidence on how discrimination and violence victimization contribute to psychosocial health problems and HIV risk among MSM and TGW [211, 212]. Stigma and violence elimination programs, and interventions, in multiple sectors (e.g., healthcare, education, employment) and social campaigns to promote understanding and acceptance of LGBTQI+ people are needed. The lack of access to gender-affirmative hormone therapy and surgeries for trans people highlights the need to improve access to such services, especially in the context of the Transgender Persons (Protection of Rights) Act, 2019, of India. This act clearly places the onus on the Indian union and state governments to provide medical gender-affirmative health services to trans people in government hospitals and to offer health insurance for gender-affirmative health services [22].

Other research areas that require increased exploration include the role of family and peer support in LGBTQI+ mental health, interventions to increase support from families and communities, and programs to eliminate discrimination and promote acceptance in healthcare, educational and workplace settings [25]. Given the deleterious impacts of stigma and discrimination on mental health and access to care, and the protective effects of social support and resilience resources, studies that integrate an understanding of social-structural contexts that affect mental health are key to effective approaches to advancing LGBTQI+ health [26]. Expanding the evidence base on LGBTQI+ health will require additional investments by national and state health research funders, including targeted funding for non-HIV-specific LGBTQI+ health research for the academic sector and for government-funded and government-run health programs on HIV (National AIDS Control Program of NACO), sexual and reproductive health and mental health (under National Health Mission), and non-communicable diseases (e.g., National Program for Prevention and Control of Cancers, Diabetes and Cardiovascular Diseases and Stroke).

Finally, few studies made explicit reference to theoretical frameworks (e.g., syndemic theory [212], minority stress theory [213], structural violence [214]), that guided study design, analysis or interpretation. For one, such theories can advance research and understanding of the needs of understudied populations, such as sexual minority women, with studies also benefitting from community-based participatory methodologies and partnerships [198, 199]. The latter can advance application of theoretical frameworks that are sensitized to community-identified concerns, self-identifications, and priorities in Indian cultural contexts [199]. Several theoretical frameworks such as gender minority stress [215], gender affirmation [216] and intersectionality [217] that have been used productively in research among trans people in western countries, especially the US, appear not to be explicitly used in studies from India. Future research should include a focus on adapting existing frameworks to meaningfully address the Indian cultural context, as well as developing new indigenous frameworks for research with LGBTQI+ people in India. Future investigations should also ensure the inclusion of diverse subgroups of trans people—not solely gender binary, but also gender non-binary trans people—and portray local gender identity terms they use as well as indigenous constructions of gender identity, rather than defaulting to western terminologies, some of which do not translate well culturally or linguistically to the Indian LGBTQI+ experience [218].

### Strengths and limitations

This scoping review should be understood in the context of study limitations. First, we limited searches to English-language texts and those included in major academic databases; however, we are not aware of Indian native language-based academic journals, given that academics and researchers largely publish in English. Second, we did not conduct quality assessments of individual studies as this is outside the purview of a scoping review; we aimed to map the field of available research, and research gaps, rather than answer a specific research question [28]. Third, we limited our review to peer-reviewed articles, for which we identified a substantial number of sources. Future scoping reviews that include grey literature from across the vast geography and cultures of India may help to broaden our understanding of the landscape of research and gaps in regard to LGBTQI+ health; this is particularly the case given the concentration of studies identified among a minority of Indian states, and conducted almost exclusively in urban areas.

## CONCLUSION

This scoping review identified key research gaps on LGBTQI+ health in India, with investigations largely limited to HIV-related issues, MSM and TGW populations, and urban study sites. This underscores the need for expanding health research in India to address the broad spectrum of LGBTQI+ people’s lives, specifically in moving beyond HIV-focused research to address mental health and non-communicable diseases, as well. Future research should address the extensive gender gap in LGBTQI+ health research in India by focusing on health needs and healthcare experiences of lesbian and bisexual women. The broader spectrum of transgender and gender nonbinary people also merits increased focus, including studies on health needs and gaps with transmasculine people. Finally, it is crucial to include sexual orientation and gender identity in national health surveys and to provide disaggregated data among LGBTQI+ subpopulations so that extant inequalities between heterosexual and cisgender people, and within LGBTQI+ people, can be documented [219]. Large-scale government-supported national health surveys among LGBTQI+ people provide a unique opportunity to document and explain health inequalities, and to identify potential solutions [220]. Strategies to enhance health research among LGBTQI+ people in India include developing a national LGBTQI+ health research agenda, providing dedicated LGBTQI+ health research funding from various government bodies, investing in the training of researchers and new investigators to competently conduct LGBTQI+ health research, and investing in improving the research and service provision capacities of community-based organizations that already bear the onus of serving a substantial number of LGBTQI+ people who are otherwise underserved by government-funded healthcare systems.

## Supporting information

S1 Appendix - Sample search strings

## Data Availability

All data underlying our findings are reported in the manuscript.

## Author Contributions

**Conceptualization:** Venkatesan Chakrapani, Peter A. Newman.

**Data curation:** Murali Shunmugam, Suchon Tepjan.

**Formal analysis:** Murali Shunmugam, Shruta Rawat, Biji R. Mohan, Venkatesan Chakrapani.

**Funding Acquisition:** Peter A. Newman, Venkatesan Chakrapani.

**Investigation:** Venkatesan Chakrapani, Peter A. Newman, Murali Shunmugam, Dicky Baruah, Biji R. Mohan.

**Methodology:** Peter A. Newman, Venkatesan Chakrapani.

**Project administration:** Murali Shunmugam, Shruta Rawat, Suchon Tepjan.

**Resources:** Suchon Tepjan.

**Software:** Suchon Tepjan, Murali Shunmugam, Dicky Baruah.

**Supervision:** Venkatesan Chakrapani, Peter A. Newman.

**Validation:** Venkatesan Chakrapani, Peter A. Newman.

**Visualization:** Murali Shunmugam.

**Writing – original draft:** Venkatesan Chakrapani, Murali Shunmugam, Shruta Rawat, Biji R. Mohan.

**Writing – review & editing:** Venkatesan Chakrapani, Peter A. Newman.

## Acknowledgements

We would like to thank Luke Reid, JD, PhD (cand.), Research Fellow, University of Toronto Factor-Inwentash Faculty of Social Work, for assistance in implementing the literature search. This review was supported by the Social Sciences and Humanities Research Council of Canada (Partnership Grant, 895–2019-1020 [MFARR-Asia]; PI: Newman). Dr. Venkatesan Chakrapani was supported in part by the DBT/Wellcome Trust India Alliance Senior Fellowship (IA/CPHS/16/1/502667).

