## Supplementary material for "A scoping review of lesbian, gay, bisexual, transgender, queer, and intersex (LGBTQI+) people’s health in India": S1 Appendix - Sample search strings

**S1 Appendix.** Sample search string for ProQuest database

AB,TI(India or “south asia” or “southeast asia”) AND (AB,TI(Bicurious or bisexual or “Kothi” OR “panthi” OR “double-decker”) OR SU(bisexuality) OR AB,TI(bisexuality or "cross sex" or bisexuals or crossgender or F2M or female-to-male or gay or gays or "gender change" or "gender dysphoria" or "gender identity" or "gender queer" or "gender reassign" or "gender transform" or "gender transition" or genderqueer or GLB or GLBQ or GLBs or GLBT or GLBTQ or heteroflexible or homosexual or homosexualities) OR SU(homosexuality) OR AB,TI(homosexuality or homosexuals or intersex or lesbian or lesbianism or lesbians or lesbigay or LGB or LGBQ or LGBS or LGBT or M2F or male-to-female or "men who have sex with men" or msm or queer or "same gender loving" or "same sex attracted" or "same sex couple" or "same sex couples" or "same sex relations" or "sex change" or "sex reassign" or "sex reversal" or "sex transform" or "sex transition" or "sexual and gender minorities" or "sexual and gender minority" or "sexual identity" or "sexual minorities" or "sexual minority" or "sexual orientation" or "sexual preference" or "trans female" or "trans male" or "trans man" or "trans men" or "trans people" or "trans person" or "trans woman" or trans-sexuality or transexual or transgender or transgendered or transgenders or transsexual) OR SU(transsexualism) or AB,TI(transsexualism or transsexuality or transsexuals or transvestite or "women loving women" or "women who have sex with women" or wsw) NOT(AB,TI("laparoscopic gastric bypass" or markov state model or multiple source method)))
